# Emergence and Epidemiology of Dominant Variants of Human Metapneumovirus in the United States between 2016 and 2021

**DOI:** 10.1101/2025.08.31.25334797

**Authors:** Lora Lee Pless, Lambodar Damodaran, Ray Pomponio, Rose Patrick, Marissa Pacey Griffith, Sara Walters, Kady D. Waggle, Atalia Pleskovitch, Vatsala Rangachar Srinivasa, Cole A. Varela, Lee H. Harrison, John P. Barton, Louise H. Moncla, Marian G. Michaels, John V. Williams, Anna F. Wang-Erickson

## Abstract

Human metapneumovirus (HMPV) causes acute respiratory disease worldwide and is the second leading cause of lower respiratory infection and hospitalization in young children in the US. There is no licensed vaccine or therapeutic. HMPV mutates rapidly; however, the specific genomic features that explain strain dominance remain undefined because there is limited routine genomic surveillance of HMPV.

We analyzed prospectively collected nasal specimens and medical data from 8,000 pediatric acute respiratory infection cases and sequenced 219 HMPV whole genomes from Pittsburgh, PA between 2016-2021. Only A2, B1, and B2 subgroups were detected; the dominant subgroup varied between seasons. Variants with an in-frame 111- or 180-nucleotide (nt) insertion that nearly duplicates the preceding flanking region in the 660-nt G gene (encodes the attachment protein) were the predominant A2 viruses detected by 2016-17. Among B2 viruses, variants with smaller in-frame insertions in the same location of the G gene became dominant by 2017-18. Each insertion length formed a distinct phylogenetic clade. The insertions are in the ectodomain and contain positively-charged residues or predicted O-glycosylation sites. Epidemiological analysis revealed that HMPV infection was independently associated with age, insurance type, and comorbidities. Elevated disease severity was independently associated with age and comorbidities, though not with HMPV subgroup. To our knowledge, in the US, this is the earliest detection of the A2 insertion variants and the first report of the B2 insertion variants. It is the largest population-based genomic HMPV study that provides a detailed phylodynamics and epidemiological analysis of prospectively collected clinical specimens.

**IMPORTANCE:** Human metapneumovirus (HMPV) is a leading cause of lung infection and pediatric hospitalizations worldwide for which there is no licensed vaccine or therapeutic. Because HMPV mutates rapidly, understanding which mutations enhance its ability to multiply and spread is important for the development of interventions and treatments. We prospectively collected patient data and nasal specimens from children with symptoms of acute respiratory illness. The predominant A2 and B2 HMPV variants circulating in the population contained insertions in the attachment protein, which suggests that these insertions may be advantageous to the virus. Furthermore, our analysis suggests that age, insurance type, and underlying health conditions were associated with HMPV infection. Age and underlying health conditions were associated with elevated HMPV disease severity, whereas HMPV subgroup was not. This large HMPV genomic epidemiological study provides insight into patient factors associated with disease and the emergence of the dominant variants in the US.

## INTRODUCTION

Human metapneumovirus (HMPV) is a significant global cause of acute respiratory disease in children and adults. Discovered in 2001, HMPV is the second leading cause of pediatric lower respiratory infection and hospitalization, costing $277M per year in the US alone (1–3). Globally, HMPV was estimated to be associated with 14.2 million acute lower respiratory cases and 643,000 hospitalizations in children under 5 years in 2018 (4). As there is no licensed vaccine or antiviral, most individuals are infected by age 5 years (5, 6). Moreover, reinfections often cause disease in older children and adults that can lead to hospitalization, especially in older adults and those with underlying comorbidities (7, 8). Because asymptomatic HPMV infection is rare, infection almost always causes distressing symptoms, including fever, rhinorrhea, cough, wheeze, otitis media, and pneumonia (2, 8, 9).

HMPV is a single stranded negative-sense RNA virus that has a 13.3 kb genome with 8 genes encoding 9 proteins (10). HMPV lineages are organized into four genotypic subgroups: A1, A2, B1, and B2 (11, 12). However, the A1 subgroup has not been detected since 2006 and may be extinct (13, 14). The virus continues to evolve, evidenced by the emergence of two A2 variants, initially detected in Japan and Spain, that harbor insertions that are near-duplications of 111 and 180 nucleotides (nt) in the G gene, which encodes the surface protein that attaches to host cells during infection (15–17). These variants have since been reported in other countries (14, 18, 19). The 111-nt variant became the dominant A2 strain circulating in Yokohama, Japan by 2018 and in Barcelona, Spain by 2021 (20, 21).

Understanding how HMPV is evolving and the predominant variants circulating in the population is critical to the development and assessment of effective future vaccines and therapeutics. However, HMPV is not included on most point-of-care rapid tests like those available for other respiratory viruses like influenza or SARS-CoV-2 (22). Clinical testing for HMPV in hospitals is not routinely ordered for all patients presenting with acute respiratory illness (ARI), since HMPV identification would not change the treatment plan as there is no targeted antiviral for HMPV (22). This limits the availability of systematically collected clinical specimens and the ability to do routine genomic surveillance to monitor how HMPV is evolving. Furthermore, sequencing the full-length HMPV genome has been historically difficult to due to regions that are prone to drop out, though recent advancements in sequencing methods optimized for HMPV have greatly improved whole genome sequencing efficiency and quality (23, 24). Consequently, studies tend to use convenience residual patient specimens, and there are few genomic epidemiological studies of HMPV (14, 19, 21, 25).

We therefore conducted the largest prospective population-based genomic epidemiological study to sequence and analyze 219 full-length HMPV genomes from pediatric specimens with data from nearly 8,000 ARI patient medical records between 2016-2021 in Pittsburgh, Pennsylvania. Because little is known about the emergence of G insertion variants in the US, we identified mutations in the G gene, assessed frequency changes over time, and described the evolutionary relationships of these variants in both local and global geographic contexts using publicly available whole genome sequences. We also identified patient factors associated with HMPV infection and disease severity.

## METHODS

### Study participants and data collection

Nasal swab specimens were collected from pediatric patients <18 years with acute respiratory illness (ARI) symptoms enrolled in inpatient, emergency department, and outpatient settings during December 1, 2016-August 31, 2021 in Pittsburgh, PA as previously described (26). Briefly, eligibility criteria included having at least one qualifying ARI sign or symptom and illness duration of <14 days. Inpatients were eligible if enrolled within 48 hours of admission. Children were excluded if they were previously enrolled in this study <14 days before the current visit or hospitalization, hospitalized <5 days after a previous hospitalization, hospitalized for a known non-respiratory cause, or had fever and neutropenia from chemotherapy. Parents or legal guardians of eligible children gave informed consent before data collection via a standardized parent / guardian interview, medical chart review, and collection of nasal specimens.

As previously described, nasal specimens were collected using flocked swabs, which were placed in universal transport medium and stored at 4°C for up to 72 hours until media was aliquoted and stored at −80°C. Reverse transcriptase quantitative PCR (RT-qPCR) was used to detect HMPV, respiratory syncytial virus (RSV), influenza, human parainfluenza 1-3 (PIV), rhinovirus / enterovirus, adenovirus, and SARS-CoV-2 (starting in 2020) (26). The study was approved by the University of Pittsburgh Institutional Review Board (STUDY19070206). This study followed the Strengthening the Reporting of Observational Studies in Epidemiology (STROBE) reporting guideline.

### Statistical Analyses

Descriptive statistics were used to summarize the demographic and clinical characteristics of acute respiratory illness (ARI) cases over the entire cohort with known HMPV status, as well as a sub-cohort with subtyped HMPV samples. An infection was defined as a positive HMPV test result. Prevalence was defined as the number of HMPV infections during the study period per total population of unique patients, and 95% confidence intervals (CIs) were estimated by bootstrap percentiles based on 1,000 samples of the patient cohort. Demographic characteristics included age, sex, race/ethnicity, and insurance type. Clinical characteristics included the presence of pre-existing conditions and viral pathogens detected. In addition to HMPV, detectable viral pathogens included influenza, para-influenza, RSV, rhinovirus, and adenovirus.

Using all ARI cases, we assessed the effect of various factors on HMPV infection status, treated as a dichotomous outcome. We reported unadjusted and adjusted odds ratios (ORs) for each factor from generalized estimating equations (GEEs), considering repeated enrollments for the same child as clustered observations by specifying an exchangeable correlation structure.

Focusing on HMPV cases only with available subgroup information, we assessed the effects of both subgroup and A2-insertion size on illness severity while adjusting for select demographic and clinical factors. Illness severity was treated as a five-level ordinal outcome and modeled using proportional odds regression. Outcome levels were as follows, in increasing order of severity: Routine ED/Clinic discharge; Admission to hospital without oxygen support; Admission to hospital with standard supplemental oxygen; Admission to ICU without invasive mechanical ventilation, extracorporeal membrane oxygenation (ECMO), or death; Admission to ICU with invasive mechanical ventilation, ECMO, or death. Proportional odds regression has been shown to improve statistical power beyond simple logistic regression in observational studies with discrete clinical endpoints, such as hospital outcomes (27). Data analyses and statistical modeling were performed using R version 4.4.1 (28).

### HMPV whole genome sequencing and assembly

#### RNA extraction and RT-qPCR

Nucleic acid was extracted from 200 µl transport media and eluted in 90 µl elution solution using the 5x MagMAX-96 Viral RNA or Viral/Pathogen Nucleic Acid kit (Applied Biosystems) and MagMAX Express 96 (Applied Biosystems) or King Fisher Apex (Thermo Fisher Scientific) liquid handlers. HMPV detection was determined by RT-qPCR with the AgPath-ID One-Step RT-PCR kit (Applied Biosystems) using the following thermocycling conditions: reverse transcription at 50°C for 30 min, reverse transcriptase inactivation/initial denaturation at 95°C for 10 min, followed by 45 cycles of 95°C for 15 min and 60°C for 30 sec.

#### cDNA synthesis and genome amplification

HMPV genomes were amplified using a tiled, overlapping amplicon-based method optimized for whole genome sequencing of HMPV(24). SuperScript IV VILO Master Mix (Thermo Fisher Scientific) was used to generate cDNA from 4 µl extracted RNA per 20 µl reaction. Platinum SuperFi PCR Master Mix (Thermo Fisher Scientific) was used to amplify each of the four amplicons that spanned the entire genome in separate 25 µl reactions and 5 µl cDNA, using the primers listed in **Table S1**. The thermocycling conditions were as follows: initial denaturation at 98°C for 30 sec; followed by 44 cycles of denaturing at 98°C for 10 sec, annealing at 60°C for 20 sec, and extension at 72°C for 2 min 10 sec; and a final extension at 72°C for 5 min. PCR products were assessed using TapeStation 4200 and the D5000 ScreenTape and reagents (Agilent).

#### Sample pooling and library preparation

The four amplicons per sample were pooled together into a single tube at approximate equimolar concentration and purified using standard Ampure XP beads (Beckman Coulter). Sequencing libraries were prepared using Illumina DNA Prep kit and unique 10 bp dual indices with a target insert size of 280 bp. Samples were sequenced on the Illumina NovaSeq X Plus, producing 2×151 bp paired-end reads.

#### Bioinformatic quality control analyses

Raw reads were demultiplexed using bcl2fastq (Illumina), adapter sequences were trimmed, and data quality control was assessed using fastp (29). Reads were classified by species using our customized reference database containing publicly available viral genomes from NCBI Virus (date accessed: March 2023) with Kraken 2 (30), followed by removal of reads mapping to the human genome (GRCh38/hg38). Next, *de novo* genome assembly was performed using rnaviralSPAdes (31) per sample, incorporating BLAST to identify the best matching viral genome from our customized viral genome database. Contigs were aligned to the best matching viral genome, and scaffolds were assembled into genomes using nucmer (32); nucleotide depth was computed using bowtie2 and bedtools (33, 34). Samples passed our quality control criteria if ≥95% of the genome was covered with at least 10× depth; the median coverage depth for all genomes was 30,433× (range, 6,662-108,777) (bedtools (34) and CheckV (35)). Protein annotations were determined using Prokka (36) and visual inspection.

### Sequence analyses

The HMPV subgroup (A1, A2, B1, or B2) was determined by comparing each sample to reference viral genome sequences using BLAST (A1 subgroup: 00-1 [NC_039199]; A2 subgroup: NL/00/17 [FJ168779]; B1 subgroup: NL/1/99 [AY525843]; and B2 subgroup: NL/94/01[FJ168778]). MAFFT v7.49 (37) was used to make multiple sequence alignments, which were subsequently inspected using Geneious Prime 2025.0.3 (www.geneious.com). A few suspected sequencing errors or assembly anomalies were removed from the final genome sequence as noted in **Table S2**. G gene insertions were detected by sequence alignment comparison to the appropriate subgroup reference sequence lacking insertions, listed above. Sanger sequencing was performed as previously described (16) to confirm our *de novo* genome assembly methods could accurately detect the presence or absence of G gene insertions using a representative sample of 50-100% of each insertion size.

### Phylodynamic analysis

A Bayesian phylogenetic modelling approach was used to characterize the transmission of HMPV in Pittsburgh locally and within the global context. Whole genome sequences for viruses collected in Pittsburgh from February 2, 2017 to April 20, 2020 were organized into subgroups A2, B1, or B2. Global contextual sequence data and associated metadata for location and date of isolation for all available HMPV whole genome sequences collected from humans in the NCBI Virus database (date accessed: August 13, 2024) (38). Data was filtered to include only complete genome sequences with associated collection year and geographic location at the country level available after January 1, 2010 **(Figure S1)**. Multiple sequence alignment was performed for all collected sequence data separately for each subgroup using MAFFT v7.2 and alignments were visually inspected using AliView v1.28 (37, 39). Because some global sequences did not have metadata for the subgroup of the sample, we performed a maximum likelihood phylogenetic reconstruction using FastTree v2.1.11, and classified sequences into global subgroup datasets based on proximity to sequences with a subgroup label in a constituent clade (40). G gene insertions in global sequences were identified as described above. A few global sequences that had insertion sizes resulting in frameshifts were annotated on the phylogeny as indicated in **Table S3**. We then created datasets to perform a local (Pittsburgh) and global analysis. For each local and global analysis, we generated one dataset for each subgroup as well as for all subgroups combined, resulting in 8 total datasets (Pittsburgh only—A2, B1, B2, and all subgroups combined; Pittsburgh + global—A2, B1, B2, and all subgroups combined). The resulting datasets had the following numbers of sequences per dataset: Pittsburgh only: A2 = 91, B1 = 53, B2 = 75, total = 219; Pittsburgh + global: A2 = 319, B1 = 114, B2 = 157, All = 590). Maximum likelihood phylogenetic reconstruction was performed for each dataset using FastTree v.2.1.11 and temporal signal was assessed using TempEst v.1.5.3 **(Figure S2, S3)** (41).

We performed phylogenetic reconstruction using BEAST v1.10.4 for all datasets with the following parameters and priors (42). For all global samples where only year of collection was provided, we set the date to the middle of the year (i.e 2016-06-01) and an uncertainty of half a year was set to cover the entire year of collection. We chose an HKY nucleotide substitution model with gamma distributed rate variation among sites and a constant size coalescent (43, 44). Based on root to tip analysis, we chose an uncorrelated lognormal relaxed clock and a uniform ucld.mean prior of 0 to 1 with mean 0.005 (45). We ran three independent MCMC chains with a chain length of 10 million states, sampling every 1,000 states, for each dataset. Runs were diagnosed using Tracer v1.7.2 to assess adequate effective sampling size across parameters, defined as an Effective Sample Score of at least 200. Tree files were combined and resampled using LogCombiner v1.10.4, discarding burn-in between 10-20%, to create a posterior sample of 10,000 trees. Maximum clade credibility (MCC) trees for each dataset were created using TreeAnnotator v1.10.4 with a posterior limit of 0.9 to annotate internal nodes. Phylogenies were visualized using ggtree v3.10.1 (46). All XMLs for BEAST analyses and scripts used for visualization of results can be found in the following Github repository: https://github.com/moncla-lab/HMPV-Pittsburgh.

### Data availability

All genome sequences generated in this study were submitted to Genbank as Bioproject PRJNA1258959 with accession numbers PV796784 - PV797002. Raw sequence reads were also uploaded to Sequence Read Archive (SRA) with accession numbers SRR33431103 – SRR33431321.

## RESULTS

### Descriptive characteristics of study population

A total of 7,982 acute respiratory illness (ARI) cases in 6,945 patients enrolled from December 1, 2016-August 31, 2021 were tested for HMPV and analyzed in this study **(Table 1)**. The overall prevalence of HMPV infection was 5.5% (379 of 6,945), 95% CI 5.1-5.9%. The median (IQR) age of ARI cases was 1.75 (0.75, 4.0) years, and 58.2% were male. The majority of ARI cases were under 5 years old (80.3%). Of the 379 HMPV cases, 112 (29.6%) were classified as co-infections where HMPV and at least one other respiratory virus was detected. Amongst co-infected HMPV cases, rhinovirus / enterovirus was the most detected other virus (18.2%), followed by adenovirus (7.8%). HMPV subgroup was determined for 249 specimens, and full-length HMPV genome sequences were obtained for 219 specimens. The characteristics of the HMPV subtyped cases were similar to those of the overall HMPV positive (HMPV+) cases (**Table 1**).

**Table 1.** Demographic and clinical characteristics by HMPV infection status.

|  | Subtyped HMPV cases, n=249<sup>a</sup> | HMPV Infection Status |  | Overall, N=7,982<sup>c</sup> | Unadjusted OR (95% CI) | Adjusted OR (95% CI) |
| --- | --- | --- | --- | --- | --- | --- |
|  |  | Negative, N=7,603 | Positive, N=379<sup>b</sup> |  |  |  |
| **Age in years, median (IQR)** | 1.33 (0.58, 3.00) | 1.83 (0.75, 4.08) | 1.42 (0.58, 3.00) | 1.75 (0.75, 4.00) | **0.92 (0.89, 0.95)** | NI |
| Age in years [min, max] | [0, 14.8] | [0.0, 17.9] | [0.0, 17.8] | [0.0, 17.9] |  |  |
| **Age group, n (%)** |  |  |  |  |  |  |
| Less than 1 year | 91 (36.5) | 2,363 (31.1) | 136 (35.9) | 2,499 (31.3) | **2.35 (1.61, 3.43)** | **2.86 (1.89, 4.35)** |
| 1 to <2 years | 54 (21.7) | 1,604 (21.1) | 81 (21.4) | 1,685 (21.1) | **2.08 (1.39, 3.11)** | **2.42 (1.58, 3.70)** |
| 2 to <3 years | 38 (15.3) | 936 (12.3) | 51 (13.5) | 987 (12.4) | **2.36 (1.52, 3.67)** | **2.63 (1.68, 4.13)** |
| 3 to <5 years | 50 (20.1) | 1,165 (15.3) | 76 (20.1) | 1,241 (15.5) | **2.80 (1.86, 4.21)** | **3.01 (1.98, 4.57)** |
| 5 years or older | 16 (6.4) | 1,535 (20.2) | 35 (9.2) | 1,570 (19.7) | REF | REF |
| **Sex, n (%)** |  |  |  |  |  |  |
| Female | 108 (43.4) | 3,175 (41.8) | 158 (41.7) | 3,333 (41.8) | 0.99 (0.80, 1.23) | 1.00 (0.80, 1.24) |
| Male | 141 (56.6) | 4,428 (58.2) | 221 (58.3) | 4,649 (58.2) | REF | REF |
| **Race and ethnicity, n (%)** |  |  |  |  |  |  |
| Black Non-Hispanic | 78 (31.7) | 2,388 (31.8) | 125 (33.3) | 2,513 (31.9) | 1.08 (0.86, 1.37) | 1.08 (0.84, 1.39) |
| White Non-Hispanic | 130 (52.8) | 4,063 (54.1) | 199 (53.1) | 4,262 (54.1) | REF | REF |
| Other Non-Hispanic<sup>d</sup> | 27 (11.0) | 740 (9.9) | 37 (9.9) | 777 (9.9) | 1.06 (0.73, 1.52) | 1.03 (0.71, 1.49) |
| Hispanic | 11 (4.5) | 315 (4.2) | 14 (3.7) | 329 (4.2) | 0.96 (0.55, 1.67) | 0.97 (0.56, 1.70) |
| **Insurance type, n (%)** |  |  |  |  |  |  |
| Private | 79 (33.1) | 2,695 (36.6) | 120 (33.0) | 2,815 (36.4) | REF | REF |
| Public | 149 (62.3) | 4,540 (61.6) | 230 (63.2) | 4,770 (61.7) | 1.11 (0.88, 1.40) | 1.05 (0.82, 1.34) |
| Both | 5 (2.1) | 41 (0.6) | 6 (1.6) | 47 (0.6) | **3.26 (1.35, 7.83)** | **3.07 (1.27, 7.43)** |
| None | 6 (2.5) | 90 (1.2) | 8 (2.2) | 98 (1.3) | **2.16 (1.02, 4.58)** | **2.22 (1.05, 4.70)** |
| **Any pre-existing condition, n (%)<sup>e</sup>** | 100 (40.3) | 2,653 (34.9) | 149 (39.4) | 2,802 (35.1) | **1.25 (1.01, 1.55)** | NI |
| **Pre-existing condition, n (%)** |  |  |  |  |  |  |
| Chronic Lung Disease<sup>e</sup> | 53 (21.4) | 1,687 (22.2) | 83 (22.0) | 1,770 (22.2) | 1.01 (0.79, 1.30) | 1.25 (0.94, 1.65) |
| Neurologic/Neuromuscular Disease<sup>g</sup> | 24 (9.7) | 500 (6.6) | 35 (9.3) | 535 (6.7) | **1.45 (1.01, 2.09)** | **1.75 (1.19, 2.56)** |
| Immunocompromised Condition | 3 (1.2) | 123 (1.6) | 4 (1.1) | 127 (1.6) | 0.69 (0.26, 1.86) | 0.84 (0.31, 2.27) |
| **HMPV cases with multiple viral detections, n (%)** | 59 (23.7) | -- | 112 (29.6) | -- | NI | NI |
| **Other viruses detected, n (%)** |  |  |  |  |  |  |
| Adenovirus | 16 (6.5) | 599 (8.1) | 29 (7.8) | 628 (8.1) | NI | NI |
| Influenza | 3 (1.2) | 644 (8.7) | 11 (2.9) | 655 (8.4) | NI | NI |
| Parainfluenza | 7 (2.8) | 695 (9.1) | 10 (2.6) | 705 (8.8) | NI | NI |
| Respiratory syncytial virus | 2 (0.8) | 1,824 (24.0) | 10 (2.6) | 1,834 (23.0) | NI | NI |
| Rhinovirus / enterovirus | 36 (14.5) | 3,075 (40.5) | 69 (18.2) | 3,144 (39.4) | NI | NI |

### Factors associated with HMPV infection and elevated disease severity

Age, insurance type, and neurologic/neuromuscular disease were each associated with HMPV infection after adjusting for other characteristics **(Table 1)**. Compared to school-aged children ≥5 years, all younger age groups had at least twice increased odds of HMPV infection (e.g. infants (<1 year) AOR, 2.86; 95% CI, 1.89-4.35). Compared to children with private insurance, those with no insurance had twice higher odds of HMPV infection (AOR, 2.22; 95% CI, 1.05-4.70), and those with both public and private insurance had 3 times higher odds of infection (AOR, 3.07; 95% CI, 1.27-7.43). Children with neurologic/neuromuscular disease had 75% increased odds of infection compared to those with no pre-existing conditions (AOR, 1.75; 95% CI, 1.19-2.56). However, children with chronic lung disease or immunocompromised condition did not have significantly increased odds of HMPV infection **(Table 1)**.

Age, chronic lung disease, and neurologic/neuromuscular disease were each associated with elevated disease severity among subtyped HMPV cases after adjusting for other characteristics **(Table 2)**. Infants had almost 3 times higher odds of elevated disease severity compared to older children ≥3 years (AOR, 2.98; 95% CI, 1.42-6.24). Children with chronic lung disease or neurologic/neuromuscular disease had nearly 5 times higher odds of elevated disease severity (chronic lung AOR, 4.88; 95% CI, 2.39-9.97; neurologic/neuromuscular AOR, 4.80; 95% CI 2.01-11.47) **(Table 2)**.

**Table 2.**
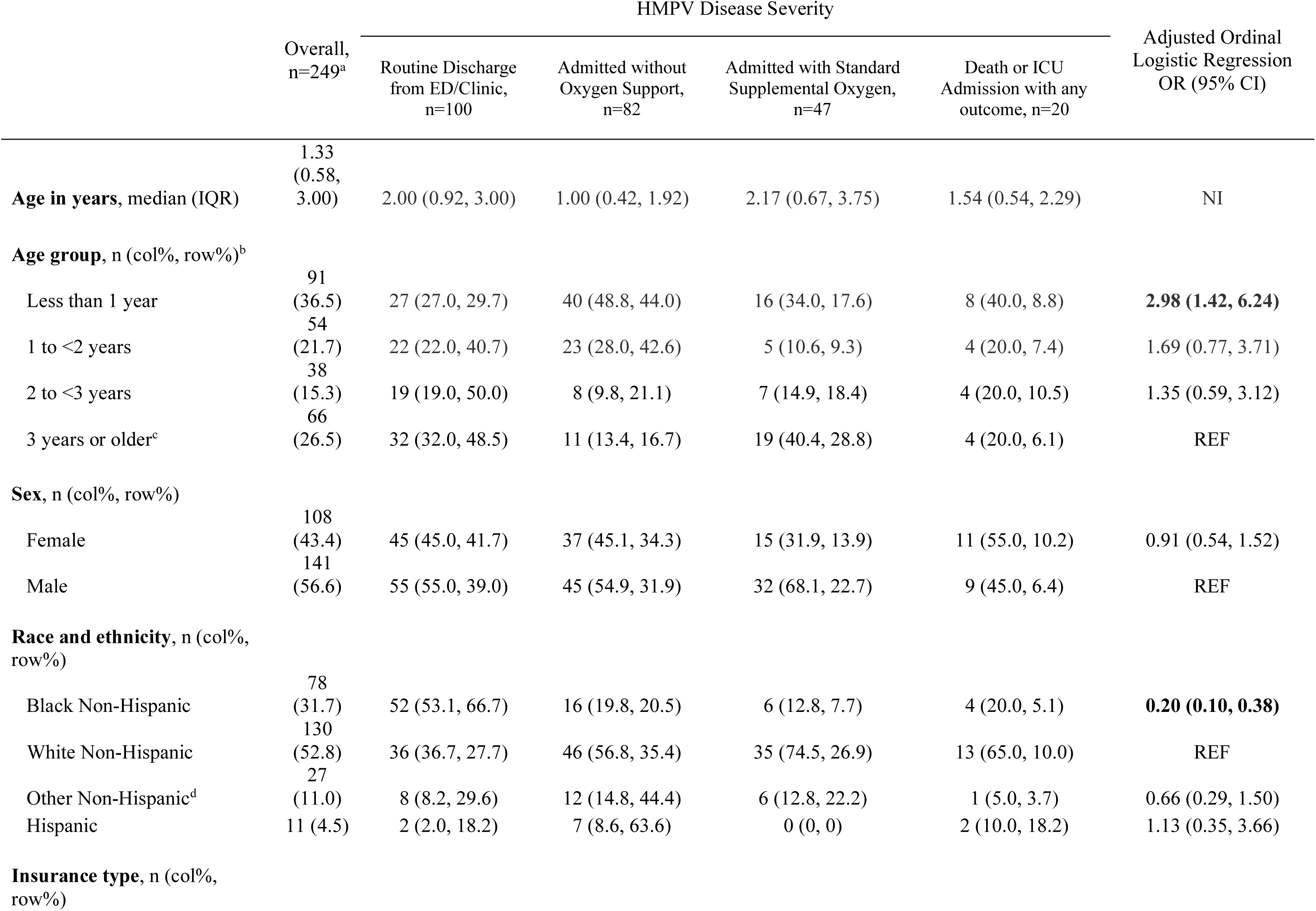
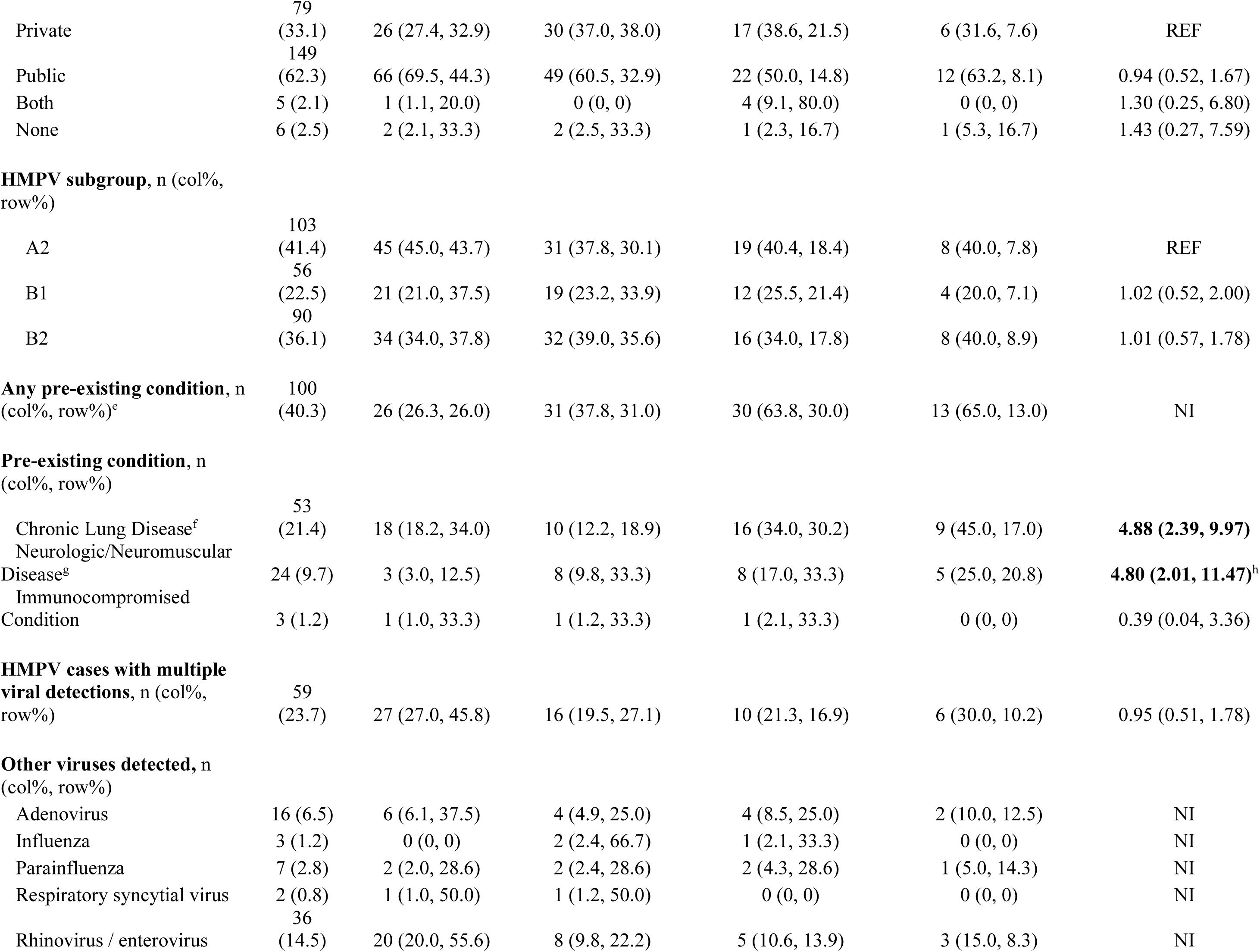
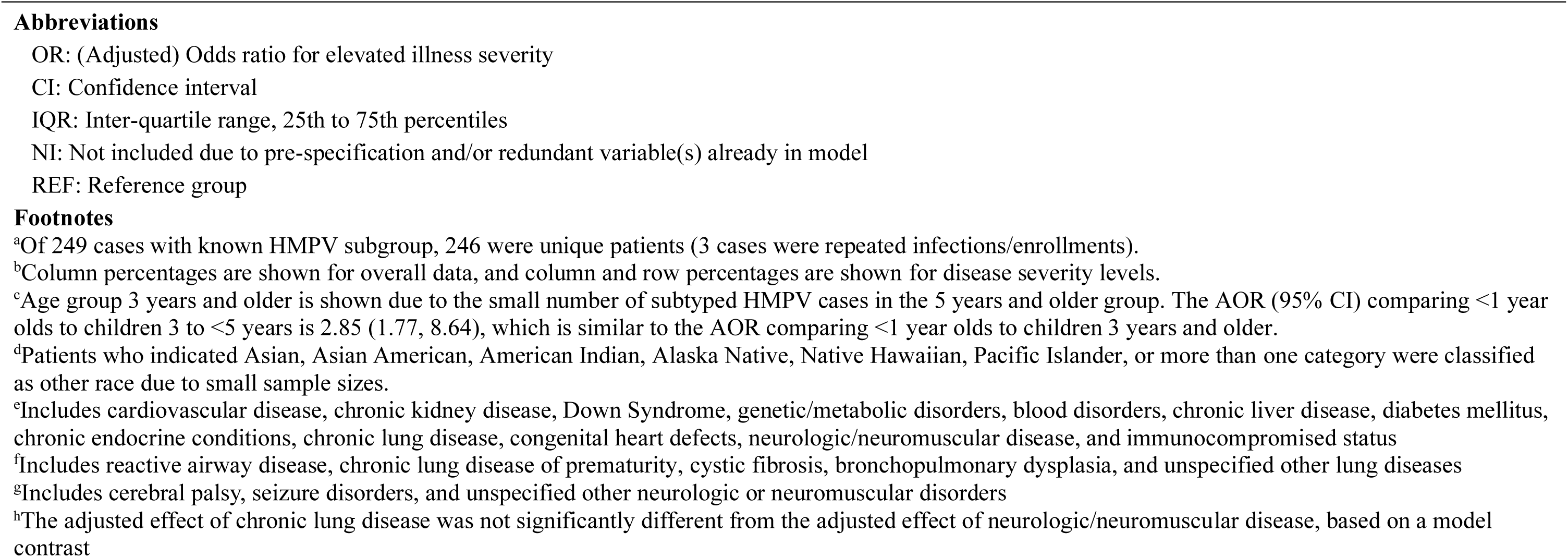
Characteristics of subtyped HMPV cases by disease severity level.

### HMPV seasonality

HMPV was more commonly detected in the spring months. HMPV consistently peaked in April every season, with test positivity reaching as high as 24.2% in April 2019 (**Figure 1A**), though this pattern was disrupted by the SARS-CoV-2 pandemic in March 2020. There were no positive HMPV samples collected from May-August 2021, except for one detection in August 2020. A2, B1, and B2 viruses co-circulate, and the dominant subgroup varied from season to season (**Figure 1B**). The A1 subgroup was not detected, consistent with previous reports that the lineage may have become extinct (14, 47).

**Figure 1.**
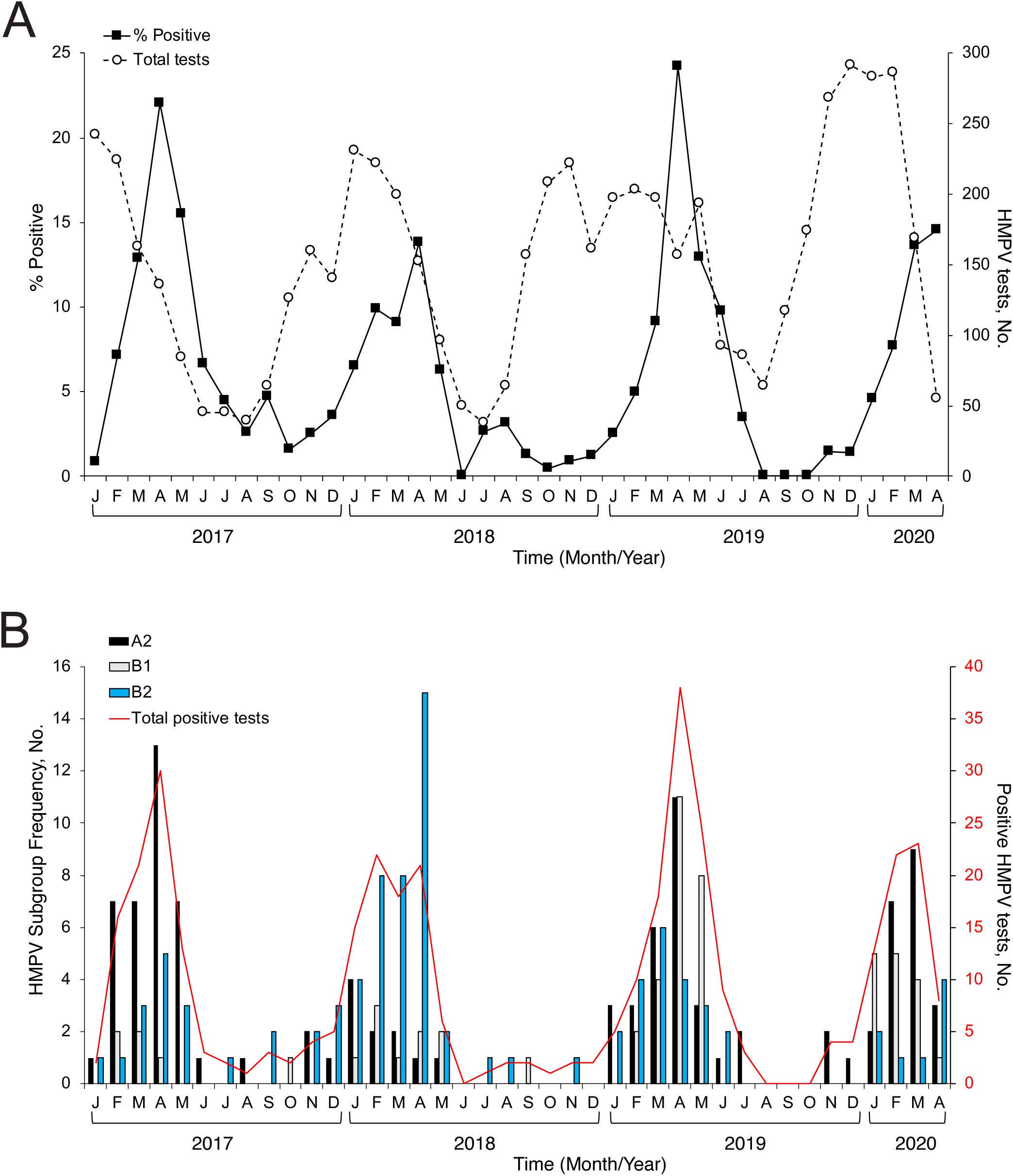
HMPV subgroup frequency and seasonality from 2017-2020. **A)** The percentage of positive HMPV tests (black squares) and total HMPV tests (open circles) are shown by month. The months December 2016 (2 positive tests) and August 2020 (1 positive test) with indeterminate subgroup are not shown. No HMPV was detected in other months after April 2020. **B)** HPMV positive specimens were sequenced to determine subgroup. Subgroup distribution by month is shown for A2 (black bars), B1 (blue bars), and B2 (gray bars). The A1 subgroup was not detected. Total number of HMPV positive tests are shown by month (red line).

After controlling for age, sex, race and ethnicity, insurance, pre-existing conditions, and co-infections, there was insufficient evidence to suggest that subgroup was associated with elevated disease severity (**Table 2**).

### A2 G gene insertion variants

Analysis of the G gene sequences revealed that nearly all A2 viruses had a 111- or 180- nucleotide (nt) insertion in the ectodomain of the G gene that nearly duplicates the upstream flanking 37 or 60 amino acid residues (**Figure 2**). There was only one detection of the classic strain with no insertion. During earlier seasons from 2017-2019, the 180-nt variant outnumbered the 111-nt variant; however, during the 2019-20 season, the proportion of the 111-nt variant rose to nearly 80% of A2 viruses detected. There was also one detection of a novel 69-nt insertion variant that was collected in 2017, but it was not detected again during the study period (**Figure 2A**). All three lengths of insertions appeared in the same location. Compared to the 111-nt variant, the 180-nt variant harbors a longer near-duplication that includes the same 111-nt region plus an additional 69 nucleotides further upstream. The 69-nt variant contains an insertion that nearly duplicates only the 69-nt region unique to the 180-nt variant (**Figure 2B, C**).

**Figure 2.**
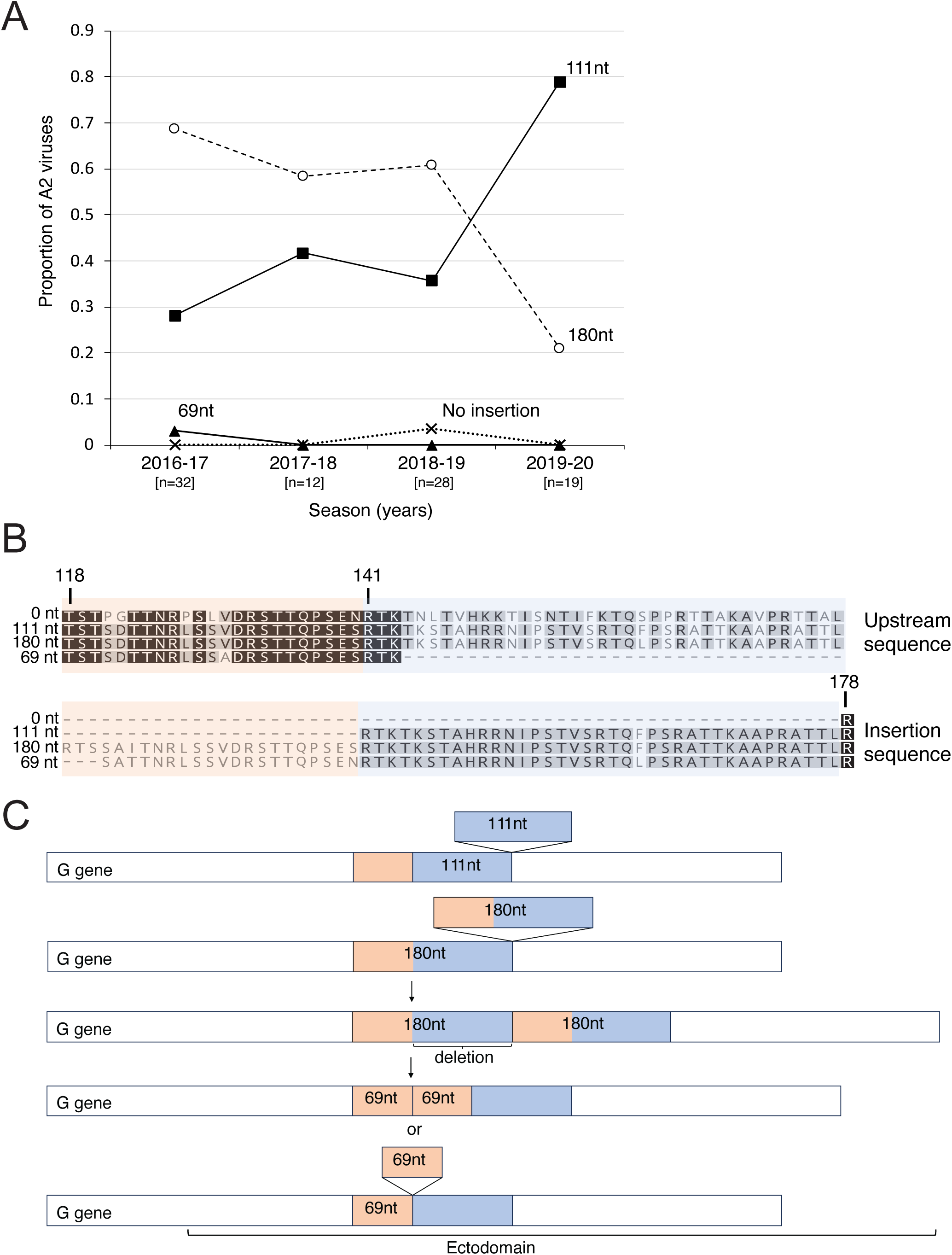
A2 variants with large insertions in the G gene. **A)** Distribution of A2 G insertion variants over time. The 180- nt variant (open circles) made up the largest proportion of A2 viruses until the 2019-20 season when the 111-nt variant (filled squares) became the most predominant detected variant. One virus with no insertion (x markers) and one virus with a 69-nt insertion (filled triangles) were detected during the study period. The total number of A2 genome sequences for each season is shown in square brackets. **B)** Excerpt of a multiple sequence alignment of G protein consensus sequences shows the insertions occur in the same location in the ectodomain. The 111-nt variant harbors an insertion that nearly duplicates the upstream 111-nt (37 amino acid) sequence (blue shading). The 180-nt variant has an insertion that nearly duplicates the same 111-nt sequence and an additional 69-nt sequence further upstream (pink shading). The 69-nt variant has a duplication of the same 69-nt sequence in the 180-nt variant. Consensus sequences were constructed by first aligning the whole genome nucleotide sequences for each variant length (111-nt variant n=39; 180-nt variant n=50), followed by extraction of the G gene nucleotide sequences. Geneious Prime was used to generate consensus G nucleotide sequences at a strict 50% threshold. The consensus sequences were translated and aligned with the 0-nt and 69-nt variant G protein sequences. Darker shaded residues indicate higher similarity. Amino acid numbering of the classic strain with no insertion is indicated using vertical lines. **C)** Cartoon drawn proportionally that illustrates that the variants harbor near-duplications of overlapping regions in the ectodomain. The 69-nt variant could have resulted from a duplication of the 69-nt sequence or a deletion of the 111-nt region from the 180-nt variant.

After controlling for age, sex, and pre-existing conditions, there was insufficient evidence to suggest that A2 insertion size was associated with elevated disease severity (**Table S4**).

### B2 G gene insertion variants

We found that many B2 HMPV specimens collected since the 2016-17 season have novel in-frame insertions of 3, 6, 9, or 12 nucleotides, encoding one to three additional amino acid residues, compared to a B2 reference sequence. The additional residues were predominantly lysine and glutamic acid. These B2 insertion sequences align to a stretch of 15 additional nucleotides in the B1 G gene that are not present in the classic B2 G gene **(Figure 3A)**. Unlike B2, no B1 viruses harbored any G gene insertions **(Figure 4B)**. During the 2016-17 season, almost half of all B2 viruses were the classic strain with no insertion; however, the classic strain was not detected in subsequent seasons during the study period. B2 insertion variants of any size were the only B2 viruses detected by the 2017-18 season, with the 3-nt variant the most commonly detected B2 variant by the 2018-2019 season **(Figure 3B)**.

**Figure 3.**
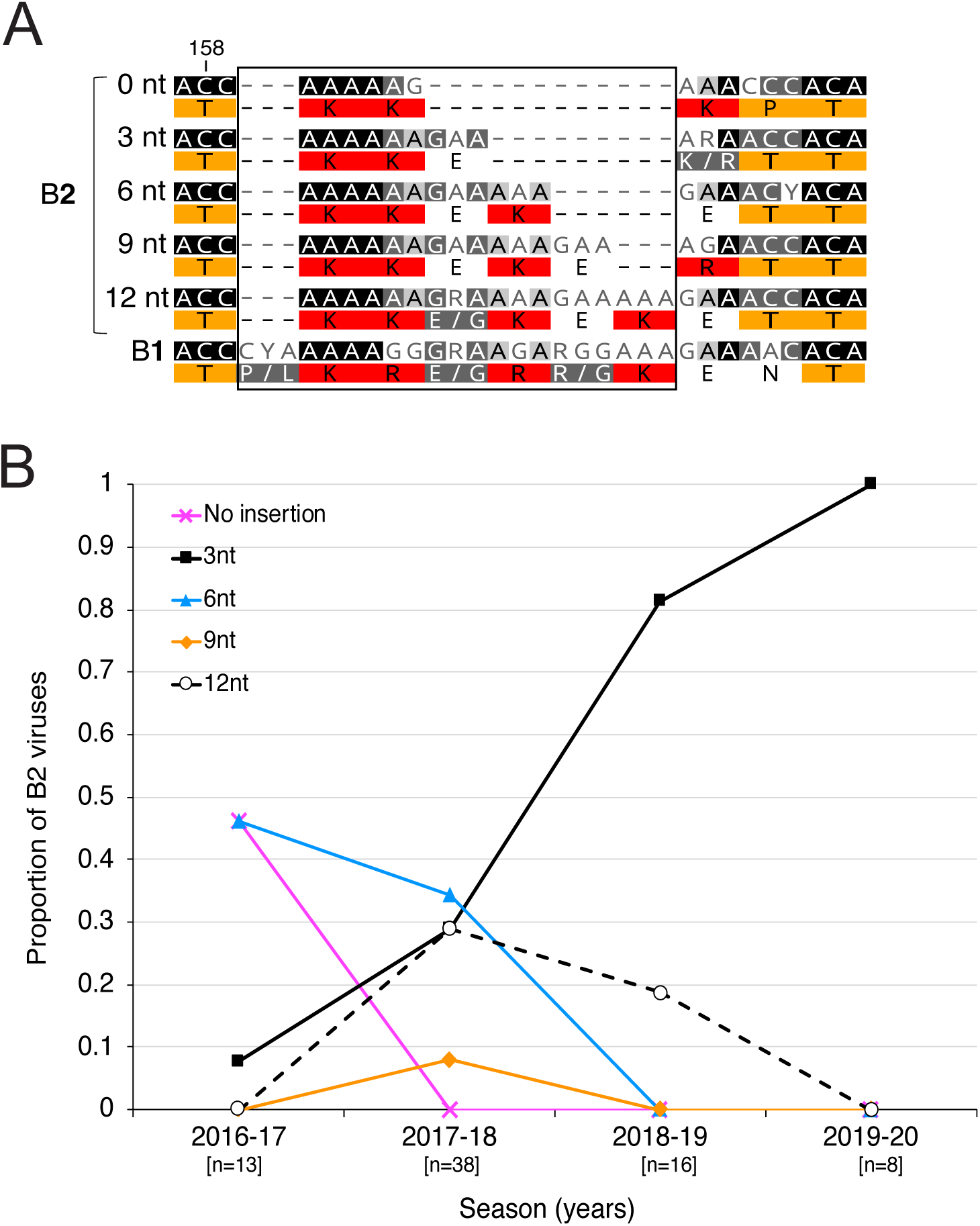
B2 variants with small insertions in the G gene. **A)** Excerpt of a multiple sequence alignment of G gene consensus sequences that shows the B2 insertions (boxed region) align to a B1 G sequence that is not present in the classic B2 strain with no insertion. Consensus sequences were constructed by first aligning the whole genome nucleotide sequences for each variant length (0-nt variant n=6; 3-nt variant n=33; 6-nt variant n=19; 9-nt variant n=3; 12-nt variant n=12), followed by extraction of the G gene nucleotide sequences. Geneious Prime was used to generate consensus G nucleotide sequences at a very strict 95% threshold. A multiple nucleotide sequence alignment was made with the consensus sequences, and the encoded amino acid residues are shown under the nucleotides. Darker shaded nucleotides indicate higher similarity. Residues are shaded using the Clustal color scheme. Amino acid numbering of the classic strain with no insertion is indicated using vertical lines. **B)** Distribution of B2 G insertion variants over time. The classic strain with no insertion (magenta line) was no longer detected after the 2016-17 season. While several lengths of insertion variants were detected during the 2017-18 season, the 3-nt variant (black squares) has become the most detected variant since 2018-19. The total number of A2 genome sequences for each season is shown in square brackets

**Figure 4.**
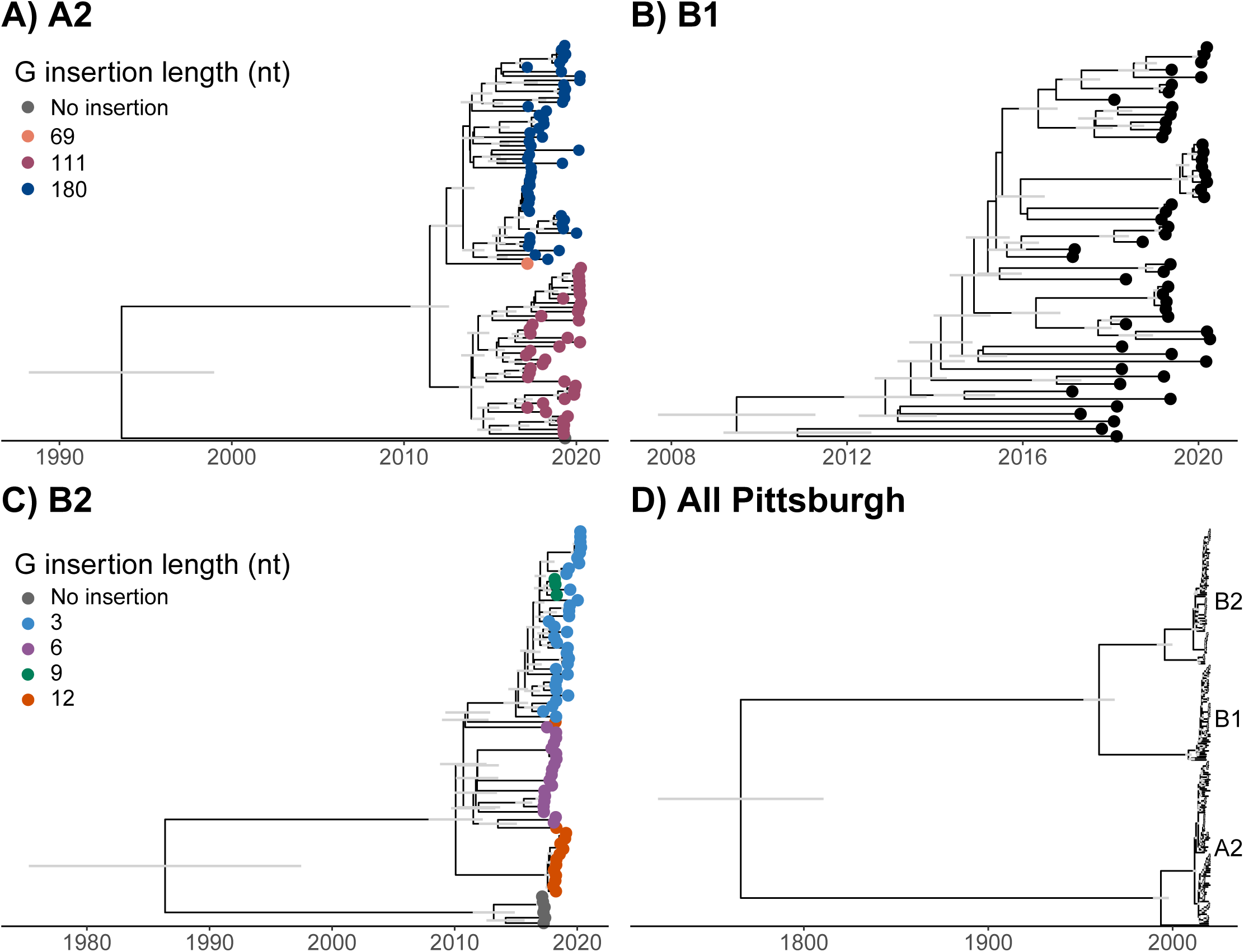
Insertions in the G gene for each genotype form distinct clades. Bayesian phylogenetic reconstructions of samples from Pittsburgh of **A)** A2 genotype **B)** B1 genotype, **C)** B2 genotype, and **D)** All Pittsburgh samples together. If insertion length data was available, tips were colored by size of the nucleotide insertion in the G gene. Grey bars indicate the 95% highest posterior density of the node height for nodes of the phylogeny that had > 90% posterior probability.

### Phylogenetic analysis

We performed a phylogenetic analysis of HMPV at the local scale (including only samples collected in Pittsburgh) and at the global scale (including Pittsburgh sequences and publicly available sequences collected globally). Insertion lengths in the A2 and B2 subgroups generally formed distinct phylogenetic clades in both the Pittsburgh (**Figure 4**) and global analyses (**Figure 5**). The clades for each insertion co-circulated across years for both subgroups. Analysis of viruses sampled globally showed higher resolution of insertion emergence and limited geographic structuring between sequences from different continents (**Figure 5**). In the global B2 analysis, the 12-nucleotide insertion emerged independently 5 times, each time evolving from viruses with the 6-nucleotide insertion **(Figure 5C)**. Pittsburgh samples were interspersed across each subgroup tree **(Figure 5, S4)**. Pittsburgh B1 and B2 viruses formed clades that were related to North American samples, whereas A2 viruses formed more distinct clades of only Pittsburgh related samples. Samples collected in Pittsburgh contributed nearly half of all available B1 and B2 sequences (46.5% and 47.8%, respectively) and nearly all available A2 180-nt variant whole genome sequences. To more accurately represent the global circulation of the virus, greater sampling is needed, especially for B1 and B2 viruses, which were sampled at a lower frequency (19.2% and 26.5% of available sequences, respectively).

**Figure 5.**
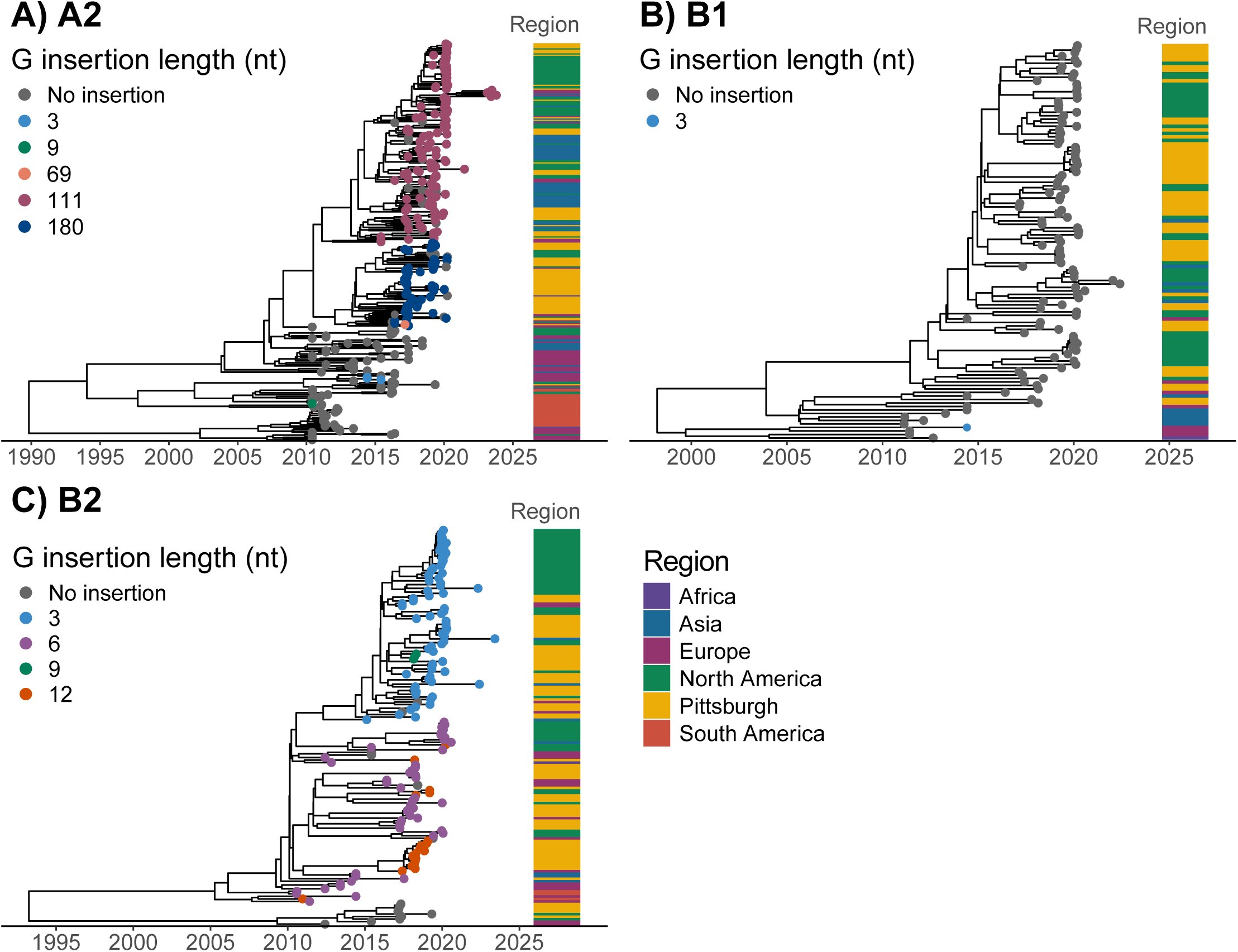
HMPV is globally heterogeneous for place of isolation across subgroups. Bayesian phylogenetic reconstructions of global samples of HPMV for the **A)** A2 genotype **B)** B1 genotype, and **C)** B2 genotype. Tips were colored by insertion length and the region where the sample was isolated. Grey bars indicate the 95% highest posterior density of the node height for nodes of the phylogeny that had > 90% posterior probability.

## DISCUSSION

This genomic epidemiological study contributed 219 full-length HMPV genome sequences collected from 2017-2020 and detected HMPV variants with insertions in the ectodomain of the G gene that became the predominant circulating strains in Pittsburgh, PA by the 2017-18 season. This suggests insertions in the G gene may confer a viral fitness advantage. Our exploratory epidemiological analysis using medical records of all subtyped HMPV samples and the entire patient cohort of nearly 8,000 ARI cases identified factors associated with HMPV infection and elevated disease severity.

Variants in the A2 subgroup harbored large (69, 111, or 180-nt) near-duplications in the ectodomain of the 660-nt G gene, whereas B2 variants had small (3, 6, 9, or 12-nt) insertions near the same region of the G gene as the A2 G insertions. No insertions were detected in B1 viral genomes. Only one A2 virus with no insertion was detected during the study period, indicating that the insertion variants had become the dominant A2 viruses before the 2016-17 season. The B2 viruses with no insertion were no longer detected after the 2016-17 season.

The A2 111- and 180-nt variants have been previously detected elsewhere (14–19). We observed that the 180-nt variant was more common than the 111-nt variant earlier in the study period, but then the 111-nt later overtook the 180-nt variant during the 2019-20 season. These results are consistent with smaller studies that reported dynamics between these two variants (14, 19–21).

The A2 G insertion sequences we detected are predicted to contain additional O-glycosylation sites, similar to the insertion sequences found in other studies (15–17). These additional O-glycosylation sites could enhance binding to host cells during infection (48). The related respiratory syncytial virus also has two sizes of G insertion variants which have become the dominant strains circulating globally, and one variant was shown to have enhanced attachment compared to wild type virus (49–51). We speculate that the 111-nt variant could have outcompeted the 180-nt variant because the excessive O-glycosylation sites in the 180-nt variant may have enhanced attachment to the point of hindering viral budding and subsequent infection, relative to the 111-nt variant. The 111-nt variant may have a moderate level of enhanced attachment, which could have resulted in a comparative advantage over the 180-nt variant.

The identification of a novel A2 69-nt insertion variant in our study raises an intriguing possibility that the insertion sequences could function as modular units, since the 180-nt insertion is the combination of the 69-nt and 111-nt sequences **(Figure 2C)**. More sampling is needed to determine whether the 69-nt variant emerged from the 180-nt variant following a deletion of the 111-nt region or as a straightforward duplication of the upstream 69-nt region.

To our knowledge, this is the first detection of B2 variants with insertions in the G gene ectodomain in the US. B2 G insertions were previously detected in Spain (21, 52), but the genetic characteristics and frequencies of different insertion lengths were not assessed. Interestingly, we found that the B2 G insertions of alternating lysine and glutamate residues align to a B1 subgroup G sequence containing a lysine-arginine-glutamate/glycine motif unique to the B1 G gene (53). Recently collected B2 variants have insertions “filling in” to copy this B1 motif with similar residues, suggesting convergent evolution between the B1 and B2 G genes. Though the function of this motif is unclear, the additional positively charged lysines in the B2 insertion could enhance viral attachment to host cells. Glutamate could form salt bridges with lysine to locally stabilize this region, since the G ectodomain appears to be unstructured (54). Though the 3-nt variant became the predominant circulating B2 virus in our sample collection, more sampling is needed to parse the dynamics between the different lengths of B2 insertion variants.

Furthermore, our whole genome phylodynamics analysis showed that the different A2 and B2 insertion variants form distinct clades and that there is limited geographical structuring, though sampling is heterogeneous across regions. Therefore, more genomic surveillance is needed to determine whether G insertions confer a viral fitness advantage that leads to continued propagation and contributes to global spread.

Multiple previous studies have evaluated whether any subgroup is associated with elevated disease severity. These studies have conflicting results where some conclude severity is associated with A1/A2 (52, 55), B1/B2 (56), or no particular subgroup (14, 57, 58). Using medical records associated with all subtyped HMPV samples, we did not detect a statistically significant difference in elevated disease severity between the subgroups. Compared to other studies, our study had a larger HMPV-positive sample size and the ability to control for variables such as age and pre-existing conditions, which we interpret to contribute the primary effects on elevated disease severity. Having either a pre-existing chronic lung or neurologic/neuromuscular disease was associated with 5 times greater odds of elevated disease severity. Though a larger proportion of patients at every level of elevated disease severity had the A2 180-nt variant compared to the 111-nt, this difference was not statistically significant at this sample size.

Our exploratory epidemiological analysis also revealed that age, insurance type, and neurologic / neuromuscular disease were associated with HMPV infection. HMPV infections are rarely detected in asymptomatic children (2, 59), and our study population was drawn from children who sought medical care for ARI symptoms. Our finding that all younger age groups have at least twice greater odds of medically-attended HMPV infection, compared to school-age children, likely stems from a higher probability of primary infection in younger children with more naïve immune systems or smaller airways that prompt parents to seek medical care. Relatedly, this study found that infants, compared to children 3 years or older, had nearly three times greater odds of elevated disease severity, possibly due to infants having smaller airways that are more easily obstructed by inflammation caused by a primary HMPV infection.

Moreover, children with no insurance or both private and public insurance had at least twice greater odds of HMPV infection compared to those with private insurance. This could be due to a complex combination of access to preventative healthcare and/or socioeconomic factors. Though investigating underlying factors was outside the scope of this study, one possible explanation is that insurance type could be related to parents having employment that does not provide adequate private health insurance. These jobs could intrinsically involve more human contact and therefore increase disease exposure to the children’s households. An association between insurance and other respiratory viral infections such as SARS-CoV-2 has also been previously published, though the effect size is much greater in this study (60). In Pennsylvania, children with severe health conditions can qualify for public insurance, allowing them to be covered by both public and private insurance plans. However, even after controlling for other covariates including comorbidities, children with both public and private insurances were still at significantly higher odds of HMPV infection compared to those with only private insurance.

Our data indicated that children with neurologic / neuromuscular conditions had 75% greater odds of HMPV infection after adjusting for all other covariates. Chronic lung disease was not significantly associated with increased odds of HMPV infection. There is almost no overlap between those with chronic lung and neurologic / neuromuscular disease in the analyzed patient cohort (only 5 have both conditions).There are few studies on HMPV and neurologic / neuromuscular conditions, and most of them are case reports or surveys evaluating whether HMPV infection causes encephalopathy or seizure (61–63). Other studies assessed HMPV disease severity and concluded that pre-existing neurologic / neuromuscular conditions were associated with more severe disease, which is consistent with our findings (1, 64, 65). However, pre-existing neurologic / neuromuscular conditions have not been well established as a factor associated with HMPV infection. A possible explanation for this association is that impaired airway clearance or pulmonary reserve could increase vulnerability to HMPV infection. This finding warrants further follow up with a larger sample size.

### Strengths and Limitations

The strengths of this study include its prospective study design with human specimens systematically collected from outpatients and inpatients with corresponding patient data from 2016-2021 that allowed investigation of disease severity and adjustment for variables such as co-infections and comorbidities. To our knowledge, this is the largest prospective population-based genomic epidemiological study of HMPV, analyzing nearly 8,000 ARI cases and contributing 219 new whole genome HMPV sequences to the existing 604 complete genomes available on NCBI Virus (66).

The study also had some limitations. Patient enrollment only captured pediatric ARI cases that presented to outpatient clinics, the emergency department, or were inpatients, which limits generalizability. Not all HMPV-positive samples could be sequenced or subtyped due to inadequate levels of intact viral RNA, which could limit the representation of some subgroups or insertion lengths. Because the G insertions are near-duplications and can be misaligned or omitted during genome assembly (52), we verified insertions detected in our samples using Sanger sequencing. However, it was not feasible to verify insertions we detected in the global sequences used in our phylogenetics analysis.

### Conclusions

This study identified patient factors associated with infection and disease severity and determined that HMPV G insertion variants were the predominant A2 and B2 viruses circulating in Pittsburgh by the 2017-18 season. Genomic surveillance and further investigation to define the role of G insertions in viral fitness are needed to better understand HMPV evolution and associated clinical outcomes.

## Data Availability

De-identified data produced in the study are available upon reasonable request to the authors.

## Author Contributions

AFWE conceived, designed, supervised, and obtained funding for the study. She drafted the initial manuscript and led data analysis and interpretation. LLP provided significant contributions to the drafting of the initial manuscript. She supervised and led experimental design and execution. LD and LM performed the phylodynamics analyses. RP performed the statistical analyses. MPG performed bioinformatics analyses. SW performed and supervised experimental work. RP, KDW, AP, VRS, CAV acquired and analyzed data. All authors critically reviewed the manuscript for important intellectual content and contributed to the acquisition, analysis, or interpretation of the data.

## Additional Contributions

The authors thank Ana Aguirre Martinez, Anna Kogos, Sreya Dey, Daniel Adesola, and Jenna Snyder for technical assistance. The authors also thank Heidi Moline and Leah Goldstein for assistance in specimen acquisition and data management.

## Funding

This work was supported by the Competitive Medical Research Fund of the UPMC Health System (AFWE), the University of Pittsburgh Clinical and Translational Science Institute Public Health Trans-Disciplinary Collaboration Pilot Grant (AFWE). AFWE was supported in part by the National Center for Advancing Translational Sciences (NCATS) National Institutes of Health (NIH) grant (KL2TR001856). The content is solely the responsibility of the authors and does not necessarily represent the official views of the NIH.

## Notes

### Competing Interest Statement

The authors have declared no competing interest.

### Author Declarations

The Institutional Review Board of The University of Pittsburgh gave ethical approval for this work (STUDY19070206).

## REFERENCES

1. Davis CR, Stockmann C, Pavia AT, Byington CL, Blaschke AJ, Hersh AL, Thorell EA, Korgenski K, Daly J, Ampofo K. 2016. Incidence, Morbidity, and Costs of Human Metapneumovirus Infection in Hospitalized Children. J Pediatric Infect Dis Soc 5:303–11.

2. Williams JV, Harris PA, Tollefson SJ, Halburnt-Rush LL, Pingsterhaus JM, Edwards KM, Wright PF, Crowe JE, Jr. 2004. Human metapneumovirus and lower respiratory tract disease in otherwise healthy infants and children. N Engl J Med 350:443–50.

3. Kahn JS. 2006. Epidemiology of human metapneumovirus. Clin Microbiol Rev 19:546–57.

4. Wang X, Li Y, Deloria-Knoll M, Madhi SA, Cohen C, Ali A, Basnet S, Bassat Q, Brooks WA, Chittaganpitch M, Echavarria M, Fasce RA, Goswami D, Hirve S, Homaira N, Howie SRC, Kotloff KL, Khuri-Bulos N, Krishnan A, Lucero MG, Lupisan S, Mira-Iglesias A, Moore DP, Moraleda C, Nunes M, Oshitani H, Owor BE, Polack FP, O’Brien KL, Rasmussen ZA, Rath BA, Salimi V, Scott JAG, Simoes EAF, Strand TA, Thea DM, Treurnicht FK, Vaccari LC, Yoshida LM, Zar HJ, Campbell H, Nair H, Respiratory Virus Global Epidemiology N. 2021. Global burden of acute lower respiratory infection associated with human metapneumovirus in children under 5 years in 2018: a systematic review and modelling study. Lancet Glob Health 9:e33–e43.

5. van den Hoogen BG, de Jong JC, Groen J, Kuiken T, de Groot R, Fouchier RA, Osterhaus AD. 2001. A newly discovered human pneumovirus isolated from young children with respiratory tract disease. Nat Med 7:719–24.

6. Schuster JE, Williams JV. 2014. Human Metapneumovirus. Microbiol Spectr 2.

7. Howard LM, Edwards KM, Zhu Y, Griffin MR, Weinberg GA, Szilagyi PG, Staat MA, Payne DC, Williams JV. 2018. Clinical Features of Human Metapneumovirus Infection in Ambulatory Children Aged 5-13 Years. J Pediatric Infect Dis Soc 7:165–168.

8. van den Hoogen BG, Osterhaus DM, Fouchier RA. 2004. Clinical impact and diagnosis of human metapneumovirus infection. Pediatr Infect Dis J 23:S25–32.

9. van den Hoogen BG, van Doornum GJ, Fockens JC, Cornelissen JJ, Beyer WE, de Groot R, Osterhaus AD, Fouchier RA. 2003. Prevalence and clinical symptoms of human metapneumovirus infection in hospitalized patients. J Infect Dis 188:1571–7.

10. van den Hoogen BG, Bestebroer TM, Osterhaus AD, Fouchier RA. 2002. Analysis of the genomic sequence of a human metapneumovirus. Virology 295:119–32.

11. van den Hoogen BG, Herfst S, Sprong L, Cane PA, Forleo-Neto E, de Swart RL, Osterhaus AD, Fouchier RA. 2004. Antigenic and genetic variability of human metapneumoviruses. Emerg Infect Dis 10:658–66.

12. Huck B, Scharf G, Neumann-Haefelin D, Puppe W, Weigl J, Falcone V. 2006. Novel human metapneumovirus sublineage. Emerg Infect Dis 12:147–50.

13. Neemuchwala A, Duvvuri VR, Marchand-Austin A, Li A, Gubbay JB. 2015. Human metapneumovirus prevalence and molecular epidemiology in respiratory outbreaks in Ontario, Canada. J Med Virol 87:269–74.

14. Groen K, van Nieuwkoop S, Meijer A, van der Veer B, van Kampen JJA, Fraaij PL, Fouchier RAM, van den Hoogen BG. 2022. Emergence and Potential Extinction of Genetic Lineages of Human Metapneumovirus between 2005 and 2021. mBio doi:10.1128/mbio.02280-22:e0228022.

15. Pinana M, Vila J, Gimferrer L, Valls M, Andres C, Codina MG, Ramon J, Martin MC, Fuentes F, Saiz R, Alcubilla P, Rodrigo C, Pumarola T, Anton A. 2017. Novel human metapneumovirus with a 180-nucleotide duplication in the G gene. Future Microbiol 12:565–571.

16. Saikusa M, Kawakami C, Nao N, Takeda M, Usuku S, Sasao T, Nishimoto K, Toyozawa T. 2017. 180-Nucleotide Duplication in the G Gene of Human metapneumovirus A2b Subgroup Strains Circulating in Yokohama City, Japan, since 2014. Front Microbiol 8:402.

17. Saikusa M, Nao N, Kawakami C, Usuku S, Sasao T, Toyozawa T, Takeda M, Okubo I. 2017. A novel 111-nucleotide duplication in the G gene of human metapneumovirus. Microbiol Immunol 61:507–512.

18. Yi L, Zou L, Peng J, Yu J, Song Y, Liang L, Guo Q, Kang M, Ke C, Song T, Lu J, Wu J. 2019. Epidemiology, evolution and transmission of human metapneumovirus in Guangzhou China, 2013-2017. Sci Rep 9:14022.

19. Goya S, Nunley EB, Longley PC, Mathis JR, Varela CG, Kim DY, Nurik M, Naccache SN, Greninger AL. 2025. Phylodynamics of human metapneumovirus and evidence for a duplication-deletion model in G gene variant evolution. J Clin Virol 180:105848.

20. Saikusa M, Nao N, Kawakami C, Usuku S, Tanaka N, Tahara M, Takeda M, Okubo I. 2019. Predominant Detection of the Subgroup A2b Human Metapneumovirus Strain with a 111-Nucleotide Duplication in the G gene in Yokohama City, Japan in 2018. Jpn J Infect Dis 72:350–352.

21. Pinana M, Gonzalez-Sanchez A, Andres C, Abanto M, Vila J, Esperalba J, Moral N, Espartosa E, Saubi N, Creus A, Codina MG, Folgueira D, Martinez-Urtaza J, Pumarola T, Anton A. 2023. The emergence, impact, and evolution of human metapneumovirus variants from 2014 to 2021 in Spain. J Infect 87:103–110.

22. Macmillan C. 2025. HMPV (Human Metapneumovirus): Your Questions Answered. https://www.yalemedicine.org/news/hmpv-your-questions-answered. Accessed June 11, 2025.

23. Groen K, van Nieuwkoop S, Bestebroer TM, Fraaij PL, Fouchier RAM, van den Hoogen BG. 2021. Whole genome sequencing of human metapneumoviruses from clinical specimens using MinION nanopore technology. Virus Res 302:198490.

24. Tulloch RL, Kok J, Carter I, Dwyer DE, Eden JS. 2021. An Amplicon-Based Approach for the Whole-Genome Sequencing of Human Metapneumovirus. Viruses 13.

25. Elias-Warren A, Bennett JC, Iwu CD, Starita LM, Stone J, Capodanno B, Prentice R, Han PD, Acker Z, Grindstaff SB, Reinhart D, Logue JK, Wolf CR, Boeckh M, Kong K, Xie H, Kim G, Greninger AL, Perofsky AC, Viboud C, Uyeki TM, Englund JA, Roychoudhury P, Chu HY. 2025. Epidemiology of Human Metapneumovirus Infection in a Community Setting, Seattle, Washington, USA. J Infect Dis 232:S78–S92.

26. Perez A, Lively JY, Curns A, Weinberg GA, Halasa NB, Staat MA, Szilagyi PG, Stewart LS, McNeal MM, Clopper B, Zhou Y, Whitaker BL, LeMasters E, Harker E, Englund JA, Klein EJ, Selvarangan R, Harrison CJ, Boom JA, Sahni LC, Michaels MG, Williams JV, Langley GE, Gerber SI, Campbell A, Hall AJ, Rha B, McMorrow M, New Vaccine Surveillance Network C. 2022. Respiratory Virus Surveillance Among Children with Acute Respiratory Illnesses - New Vaccine Surveillance Network, United States, 2016-2021. MMWR Morb Mortal Wkly Rep 71:1253–1259.

27. Ceyisakar IE, van Leeuwen N, Dippel DWJ, Steyerberg EW, Lingsma HF. 2021. Ordinal outcome analysis improves the detection of between-hospital differences in outcome. BMC Med Res Methodol 21:4.

28. R Core Team. 2024. R: A language and environment for statistical computing, R Foundation for Statistical Computing, Vienna, Austria. https://www.R-project.org/.

29. Chen S, Zhou Y, Chen Y, Gu J. 2018. fastp: an ultra-fast all-in-one FASTQ preprocessor. Bioinformatics 34:i884–i890.

30. Wood DE, Lu J, Langmead B. 2019. Improved metagenomic analysis with Kraken 2. Genome Biol 20:257.

31. Meleshko D, Hajirasouliha I, Korobeynikov A. 2021. coronaSPAdes: from biosynthetic gene clusters to RNA viral assemblies. Bioinformatics 38:1–8.

32. Marçais G, Delcher AL, Phillippy AM, Coston R, Salzberg SL, Zimin A. 2018. MUMmer4: A fast and versatile genome alignment system. PLoS Comput Biol 14:e1005944.

33. Langmead B, Salzberg SL. 2012. Fast gapped-read alignment with Bowtie 2. Nat Methods 9:357–9.

34. Quinlan AR, Hall IM. 2010. BEDTools: a flexible suite of utilities for comparing genomic features. Bioinformatics 26:841–2.

35. Nayfach S, Camargo AP, Schulz F, Eloe-Fadrosh E, Roux S, Kyrpides NC. 2021. CheckV assesses the quality and completeness of metagenome-assembled viral genomes. Nat Biotechnol 39:578–585.

36. Seemann T. 2014. Prokka: rapid prokaryotic genome annotation. Bioinformatics 30:2068–9.

37. Katoh K, Standley DM. 2013. MAFFT multiple sequence alignment software version 7: improvements in performance and usability. Mol Biol Evol 30:772–80.

38. Brister JR, Ako-Adjei D, Bao Y, Blinkova O. 2015. NCBI viral genomes resource. Nucleic Acids Res 43:D571–7.

39. Larsson A. 2014. AliView: a fast and lightweight alignment viewer and editor for large datasets. Bioinformatics 30:3276–8.

40. Price MN, Dehal PS, Arkin AP. 2010. FastTree 2--approximately maximum-likelihood trees for large alignments. PLoS One 5:e9490.

41. Rambaut A, Lam TT, Max Carvalho L, Pybus OG. 2016. Exploring the temporal structure of heterochronous sequences using TempEst (formerly Path-O-Gen). Virus Evol 2:vew007.

42. Suchard MA, Lemey P, Baele G, Ayres DL, Drummond AJ, Rambaut A. 2018. Bayesian phylogenetic and phylodynamic data integration using BEAST 1.10. Virus Evol 4:vey016.

43. Drummond AJ, Rambaut A, Shapiro B, Pybus OG. 2005. Bayesian coalescent inference of past population dynamics from molecular sequences. Mol Biol Evol 22:1185–92.

44. Yang Z. 1994. Maximum likelihood phylogenetic estimation from DNA sequences with variable rates over sites: approximate methods. J Mol Evol 39:306–14.

45. Drummond AJ, Ho SY, Phillips MJ, Rambaut A. 2006. Relaxed phylogenetics and dating with confidence. PLoS Biol 4:e88.

46. Yu G, Smith DK, Zhu H, Guan Y, Lam TT-Y. 2017. ggtree: an r package for visualization and annotation of phylogenetic trees with their covariates and other associated data. Methods in Ecology and Evolution 8:28–36.

47. Aberle JH, Aberle SW, Redlberger-Fritz M, Sandhofer MJ, Popow-Kraupp T. 2010. Human metapneumovirus subgroup changes and seasonality during epidemics. Pediatr Infect Dis J 29:1016–8.

48. Zhai X, Yuan Y, He WT, Wu Y, Shi Y, Su S, Du Q, Mao Y. 2024. Evolving roles of glycosylation in the tug-of-war between virus and host. Natl Sci Rev 11:nwae086.

49. Hotard AL, Laikhter E, Brooks K, Hartert TV, Moore ML. 2015. Functional Analysis of the 60-Nucleotide Duplication in the Respiratory Syncytial Virus Buenos Aires Strain Attachment Glycoprotein. J Virol 89:8258–66.

50. Eshaghi A, Duvvuri VR, Lai R, Nadarajah JT, Li A, Patel SN, Low DE, Gubbay JB. 2012. Genetic variability of human respiratory syncytial virus A strains circulating in Ontario: a novel genotype with a 72 nucleotide G gene duplication. PLoS One 7:e32807.

51. Trento A, Galiano M, Videla C, Carballal G, García-Barreno B, Melero JA, Palomo C. 2003. Major changes in the G protein of human respiratory syncytial virus isolates introduced by a duplication of 60 nucleotides. J Gen Virol 84:3115–3120.

52. Pinana M, Vila J, Maldonado C, Galano-Frutos JJ, Valls M, Sancho J, Nuvials FX, Andres C, Martin-Gomez MT, Esperalba J, Codina MG, Pumarola T, Anton A. 2020. Insights into immune evasion of human metapneumovirus: novel 180- and 111- nucleotide duplications within viral G gene throughout 2014-2017 seasons in Barcelona, Spain. J Clin Virol 132:104590.

53. Yang CF, Wang CK, Tollefson SJ, Lintao LD, Liem A, Chu M, Williams JV. 2013. Human metapneumovirus G protein is highly conserved within but not between genetic lineages. Arch Virol 158:1245–52.

54. Leyrat C, Paesen GC, Charleston J, Renner M, Grimes JM. 2014. Structural insights into the human metapneumovirus glycoprotein ectodomain. J Virol 88:11611–6.

55. Vicente D, Montes M, Cilla G, Perez-Yarza EG, Perez-Trallero E. 2006. Differences in clinical severity between genotype A and genotype B human metapneumovirus infection in children. Clin Infect Dis 42:e111–3.

56. Papenburg J, Hamelin ME, Ouhoummane N, Carbonneau J, Ouakki M, Raymond F, Robitaille L, Corbeil J, Caouette G, Frenette L, De Serres G, Boivin G. 2012. Comparison of risk factors for human metapneumovirus and respiratory syncytial virus disease severity in young children. J Infect Dis 206:178–89.

57. Pitoiset C, Darniot M, Huet F, Aho SL, Pothier P, Manoha C. 2010. Human metapneumovirus genotypes and severity of disease in young children (n = 100) during a 7-year study in Dijon hospital, France. J Med Virol 82:1782–9.

58. Ye H, Zhang S, Zhang K, Li Y, Chen D, Tan Y, Liang L, Liu M, Liang J, An S, Wu J, Zhu X, Li M, He Z. 2023. Epidemiology, genetic characteristics, and association with meteorological factors of human metapneumovirus infection in children in southern China: A 10-year retrospective study. Int J Infect Dis 137:40–47.

59. Edwards KM, Zhu Y, Griffin MR, Weinberg GA, Hall CB, Szilagyi PG, Staat MA, Iwane M, Prill MM, Williams JV, New Vaccine Surveillance N. 2013. Burden of human metapneumovirus infection in young children. N Engl J Med 368:633–43.

60. Wang-Erickson AF, Zhang X, Dauer K, Zerr DM, Adler A, Englund JA, Lee B, Schuster JE, Selvarangan R, Rohlfs C, Staat MA, Sahni LC, Boom JA, Balasubramani GK, Williams JV, Michaels MG. 2025. Prevalence of SARS-CoV-2 in Children Identified by Preprocedural Testing at 5 US Children’s Hospital Systems. Pediatr Infect Dis J 44:47–53.

61. Arnold JC, Singh KK, Milder E, Spector SA, Sawyer MH, Gavali S, Glaser C. 2009. Human metapneumovirus associated with central nervous system infection in children. Pediatr Infect Dis J 28:1057–60.

62. Vehapoglu A, Turel O, Uygur Sahin T, Kutlu NO, Iscan A. 2015. Clinical Significance of Human Metapneumovirus in Refractory Status Epilepticus and Encephalitis: Case Report and Review of the Literature. Case Rep Neurol Med 2015:131780.

63. Mori A, Kawano Y, Hara S, Numoto S, Kurahashi H, Okumura A. 2023. A nationwide survey of human metapneumovirus-associated encephalitis/encephalopathy in Japan. Brain Dev 45:197–204.

64. Spaeder MC, Custer JW, Bembea MM, Aganga DO, Song X, Scafidi S. 2013. A Multicenter Outcomes Analysis of Children With Severe Viral Respiratory Infection Due to Human Metapneumovirus. Pediatric Critical Care Medicine 14:268–272.

65. Hahn A, Wang W, Jaggi P, Dvorchik I, Ramilo O, Koranyi K, Mejias A. 2013. Human metapneumovirus infections are associated with severe morbidity in hospitalized children of all ages. Epidemiol Infect 141:2213–23.

66. National Library of Medicine (US) NCfBI. NCBI Virus. https://www.ncbi.nlm.nih.gov/labs/virus/vssi/#/. Accessed June 5, 2025.

