## Supplemental files for "Emergence and Epidemiology of Dominant Variants of Human Metapneumovirus in the United States between 2016 and 2021"

**Table S1. Primer sequences and expected amplicon sizes.**

| Amplicon | Primer Name | Sequence (5' → 3') | Amplicon Size (bp) |
|---|---|---|---|
| 1 | hMPV1_F | GGGACAAATAAAAATGTCTCTTCA | 4125 |
| | hMPV1_F2 | GGGGAARCATGCTATATTAAAAG | |
| | hMPV1_R | CTTCCTGTGCTRACYTTRCA | |
| 2 | hMPV2_F | ACAGCAGCRGGRATYAATGT | 4010 |
| | hMPV2_F3 | GWTCMWACATGCCRACATCTG | |
| | hMPV2_R | TAGTACTGAAYTGAGCATGYTCAG | |
| 3 | hMPV3_F | AACTGTTAACATGGAAAGATGTGATG | 3229 |
| | hMPV3_R | TAAGCTGGAACWGAWGCTG | |
| 4 | hMPV4_F | TCAATAGGGAGTCTRTGTCARGAA | 3675 |
| | hMPV4_F3 | GGTCATAAACTCAAAGAAGGTG | |
| | hMPV4_R | GRCAAAAAAACCGTATACATYC | |

**Table S2. Pittsburgh genome sequences modified due to suspected sequencing errors or assembly anomaly that led to frameshifts.**

| Sample name | Subgroup | Modification |
|---|---|---|
| EP1R-02180 | A2 | Removed 1nt insertion in the N gene that resulted in frameshift and protein truncation |
| EP2R-02946 | B1 | Removed 1nt insertion in the M gene that resulted in frameshift and protein truncation |
| EP1R-00769 | B2 | Removed 71 nt insertion in the N gene that resulted in frameshift and protein truncation |
| EP1R-02295 | B2 | Removed 1 nt insertions in the M and F genes that resulted in frameshifts and protein truncation |
| EP2R-00490 | B2 | Removed 1 ambiguous base insertion in the N gene |

**Table S3. G insertion variant annotations for global sequences with frameshifts or other anomalies in the G gene due to suspected sequencing errors.** Sequences were not modified in the phylogenetic analysis.

| **Accession** | **Subgroup** | **G insertion size** | **Reason for annotation** | **Annotation on phylogeny:** |
|---|---|---|---|---|
| OL794386 | A2 | 181 | Frameshift leads to early stop codon in the insertion | 180-nt |
| PP947671 | A2 | 104 | Has 104-nt insertion but also has upstream 16nt "deletion" with early stop codon | 0-nt |
| PP947655 | A2 | 66 | Insertion is mostly contains ambiguous bases (Ns) | 0-nt |
| OL794430 | A2 | 1 | Frameshift leads to early stop codon | 0-nt |
| KY474529 | A2 | 1 | 1-nt insertion in a string of Ns. Also a missing base (C) upstream, so no net insertion | 0-nt |
| OL794402 | B2 | 4 | Frameshift leads to early stop codon | 0-nt |
| OL794432 | B2 | 1 | Frameshift leads to early stop codon | 0-nt |
| OL794475 | B2 | 1 | Frameshift leads to early stop codon | 0-nt |
| OL794441 | B2 | 5 | Frameshift leads to early stop codon | 0-nt |

**Table S4. Characteristics of cases with A2 insertion variants by disease severity level**

| | Overall, n=89[a] | HMPV Disease Severity | | | | Adjusted Ordinal Logistic Regression OR (95% CI) |
|---|---|---|---|---|---|---|
| | | Routine Discharge from ED/Clinic, n=38 | Admitted without Oxygen Support, n=26 | Admitted with Standard Supplemental Oxygen, n=18 | Death or ICU Admission with any outcome, n=7 | |
| **Age in years**, median (IQR) | 1.33 (0.67, 2.33) | 1.13 (0.75, 2.00) | 1.00 (0.42, 1.92) | 3.17 (1.00, 4.67) | 1.67 (0.58, 2.00) | NI |
| **Age group**, n (col%, row%)[b] | | | | | | |
| Less than 1 year | 31 (34.8) | 14 (36.8, 45.2) | 10 (38.5, 32.3) | 4 (22.2, 12.9) | 3 (42.9, 9.7) | 0.52 (0.15, 1.76) |
| 1-2 years | 27 (30.3) | 13 (34.2, 48.1) | 10 (38.5, 37.0) | 3 (16.7, 11.1) | 1 (14.3, 3.7) | 0.33 (0.10, 1.07) |
| 2-3 years | 13 (14.6) | 7 (18.4, 53.8) | 3 (11.5, 23.1) | 1 (5.6, 7.7) | 2 (28.6, 15.4) | 0.33 (0.08, 1.47) |
| 3 years or older | 18 (20.2) | 4 (10.5, 22.2) | 3 (11.5, 16.7) | 10 (55.6, 55.6) | 1 (14.3, 5.6) | REF |
| **Sex**, n (col%, row%) | | | | | | |
| Female | 40 (44.9) | 20 (52.6, 50.0) | 10 (38.5, 25.0) | 5 (27.8, 12.5) | 5 (71.4, 12.5) | 1.05 (0.45, 2.47) |
| Male | 49 (55.1) | 18 (47.4, 36.7) | 16 (61.5 32.7) | 13 (72.2, 26.5) | 2 (28.6, 4.1) | REF |
| **Any pre-existing condition**, n (col%, row%)[c] | 33 (37.5) | 9 (24.3, 27.3) | 9 (34.6, 27.3) | 12 (66.7, 36.4) | 3 (42.9, 9.1) | 2.25 (0.86, 5.84) |
| **HMPV A2 insertion size**, n (col%, row%) | | | | | | |
| 111nt | 39 (43.8) | 19 (50.0, 48.7) | 10 (38.5, 25.6) | 8 (44.4, 20.5) | 2 (28.6, 5.1) | REF |
| 180nt | 50 (56.2) | 19 (50.0, 38.0) | 16 (61.5, 32.0) | 10 (55.6, 20.0) | 5 (71.4, 10.0) | 1.37 (0.62, 3.05) |

**Abbreviations**

OR: (Adjusted) Odds ratio for elevated illness severity
CI: Confidence interval
IQR: Inter-quartile range, 25th to 75th percentiles
NI: Not included due to pre-specification and/or redundant variable(s) already in model
REF: Reference group

**Footnotes**

[a]All were unique patients
[b]Column percentages are shown for overall data, and column and row percentages are shown for disease severity levels.
[c]Includes cardiovascular disease, chronic kidney disease, Down Syndrome, genetic/metabolic disorders, blood disorders, chronic liver disease, diabetes mellitus, chronic endocrine conditions, chronic lung disease, congenital heart defects, neurologic/neuromuscular disease, and immunocompromised status

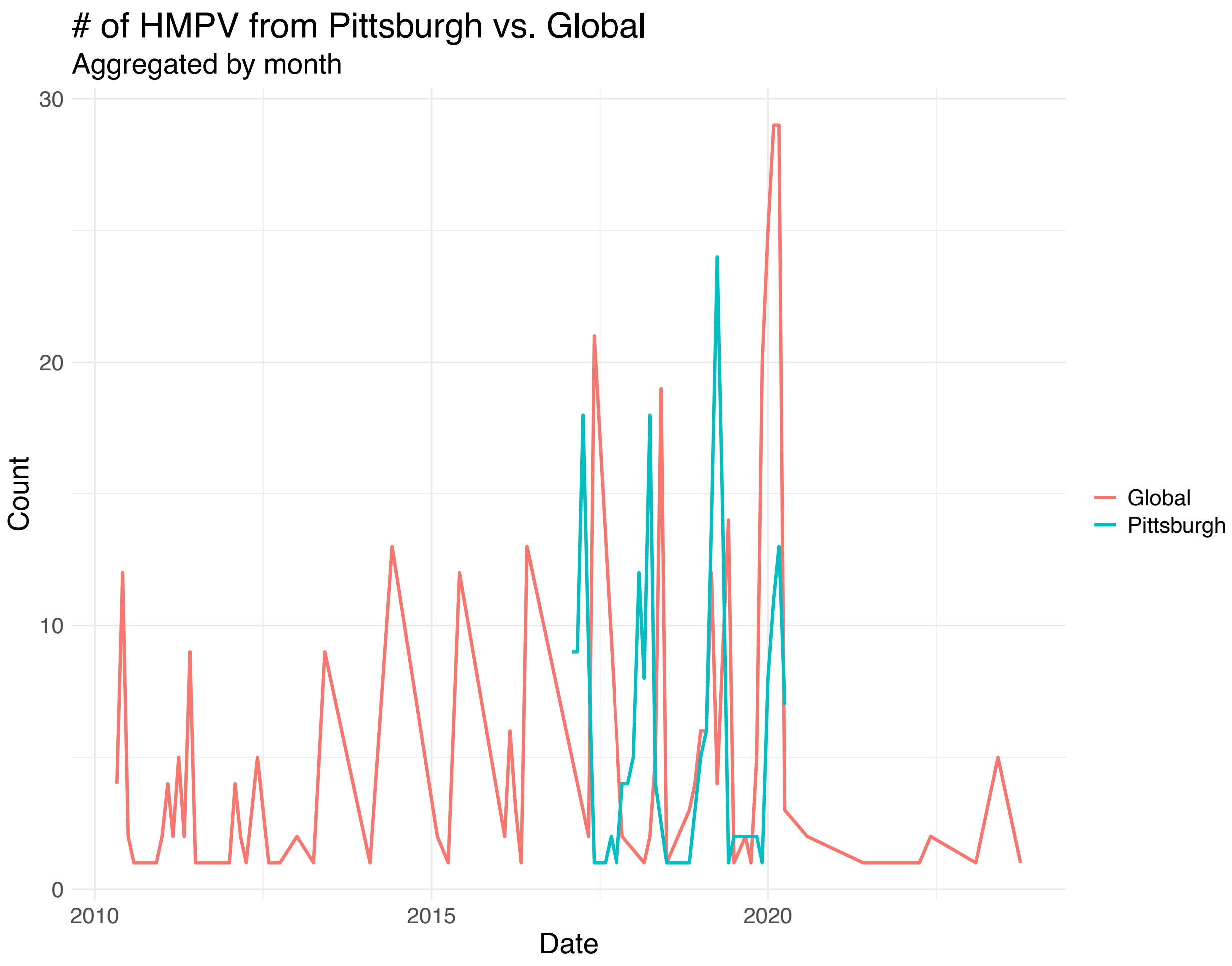


**Figure S1.** Number of HMPV full-length genome sequences from Pittsburgh generated by this study versus globally sampled sequences from NCBI Virus from January 1, 2010 to August 13, 2024.

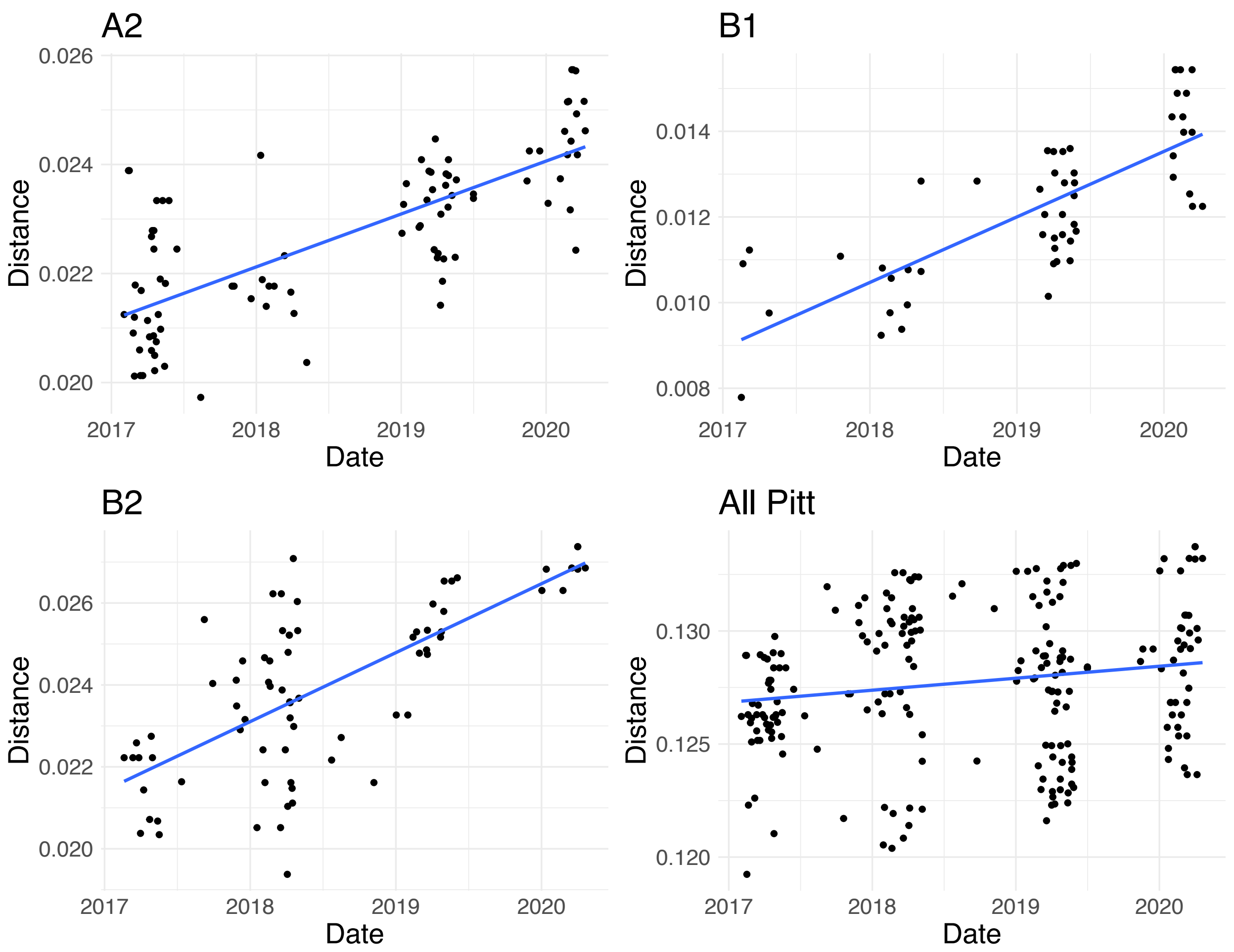


**Figure S2.** Root-to-tip regression results from TempEst v1.5.3 for Pittsburgh HMPV sequences generated in this study.

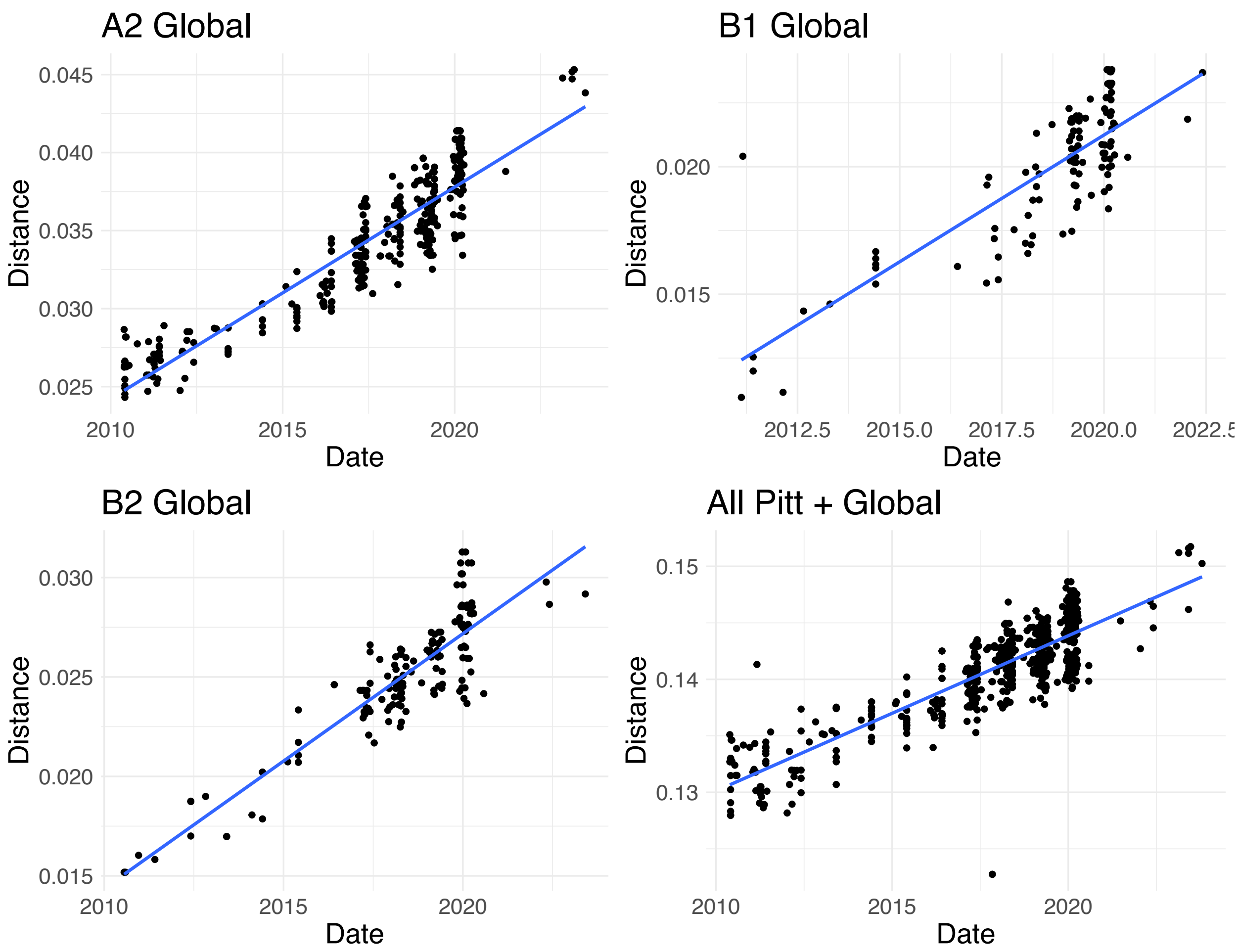


**Figure S3.** Root-to-tip regression results from TempEst v1.5.3 for global HMPV sequences from NCBI Virus.

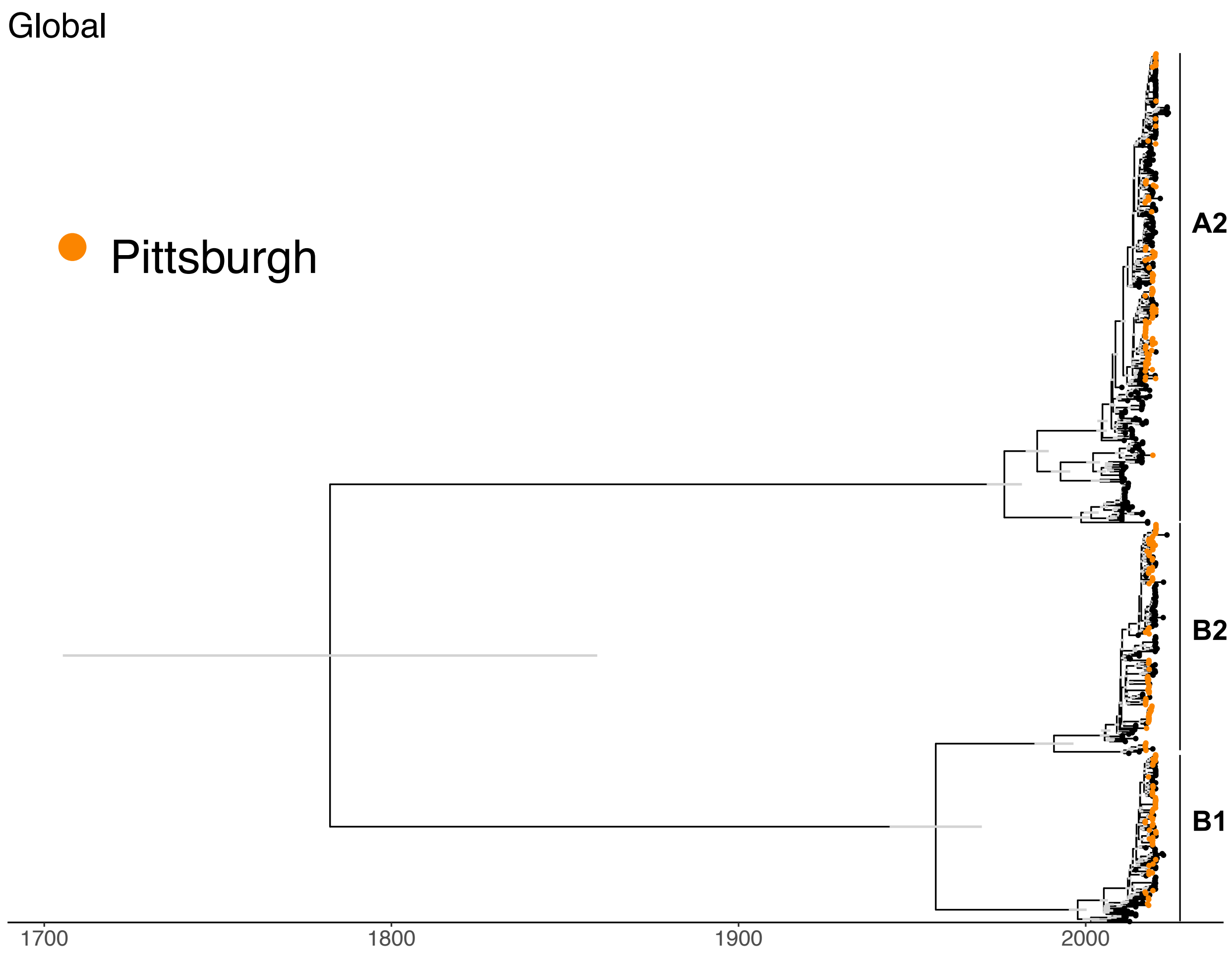


**Figure S4.** Bayesian phylogenetic reconstruction of all available whole genome sequenced samples between January 1, 2010 and August 13, 2024. Tips corresponding to samples from Pittsburgh are colored and genotypes are labeled. Grey bars indicate the 95% HPD of the node height for nodes of the phylogeny that had > 90% posterior probability.
